# Artificial Intelligence (AI)-Powered H&E Whole-Slide Image Analysis of Tertiary Lymphoid Structure (TLS) Independently Predicts Survival in Patients with Non-Small Cell Lung Cancer (NSCLC) Receiving Immunotherapy

**DOI:** 10.1101/2025.08.07.25333135

**Authors:** Leeseul Kim, Sangwon Shin, Behtash G Nezami, Borislav Alexiev, Soo Ick Cho, Sanghoon Song, Wonkyung Jung, Lee AD Cooper, Donghoon Shin, Junho Song, Taekyu Um, Liam Il-Young Chung, Young Kwang Chae

## Abstract

**Background:** Tertiary lymphoid structures (TLSs) within the tumor microenvironment have emerged as potential indicators of treatment response to immune checkpoint inhibitors (ICIs). This study aims to explore the association between artificial intelligence (AI)-powered TLS analysis and treatment outcomes in NSCLC patients treated with ICIs.

**Methods:** An AI-powered analyzer, Lunit SCOPE, was developed and trained on H&E-stained whole slide images to quantify TLS. An external cohort of 102 advanced-stage NSCLC patients was utilized to assess the predictive value of TLS.

**Results:** Of 102 patients, 30.3% (31/102) cases were found to contain TLS as determined by the AI analyzer. The AI model achieved an accuracy of 88.2% in identifying the presence of TLS. TLS presence assessed by AI analyzer was associated with significantly favorable progression-free survival (HR 0.49, 95% CI 0.29–0.82, *p* < 0.01) and overall survival (HR 0.55, 95% CI 0.33–0.92, *p* = 0.02). The presence of TLS was associated with favorable survival outcomes, independent of PD-L1 status, treatment regimen (ICI monotherapy vs. combination therapy), lines of therapy (first-line vs. second-line and beyond), and tissue harvest site (primary vs. metastatic). The TLS presence enhanced predictive performance when TLS was assessed from WSI obtained from metastatic lesions.

**Conclusions:** To our knowledge, this study is the first to develop an AI model capable of detecting the presence of TLS in WSI, with clinical validation using real-world data demonstrating its independent predictive value of survival outcomes in advanced-stage NSCLC patients receiving immunotherapy.

## INTRODUCTION

Immune checkpoint inhibitors (ICIs) have significantly transformed the therapeutic approach to non-small cell lung cancer (NSCLC)^1,2^. However, the meaningful responses to ICIs are limited to only a portion of patients^3,4^. This has led to the exploration of predictive biomarkers for immunotherapy response, such as PD-L1 and tumor mutational burden (TMB). Unfortunately, the performance of these markers is not consistently optimal, spurring ongoing research into various aspects of the tumor microenvironment (TME) as complementary indicators^5–7^.

Within the TME, tertiary lymphoid structures (TLSs), characterized as organized aggregates of immune cells at tumor sites, have emerged as a significant factor in antitumor immunity^8,9^. TLSs are associated with favorable prognosis and improved immunotherapy responses across several cancers, including NSCLC^8,10,11^. Despite their potential, challenges remain in standardizing TLS assessment due to their complex composition^8^. While multiplex immunohistochemistry (IHC) provides detailed evaluation, its cost and technical demands limit its clinical applicability. Hematoxylin and eosin (H&E) staining offers a simpler alternative but is hindered by interobserver variability^12^.

Advances in digital pathology and whole slide imaging (WSI) have enabled the integration of artificial intelligence (AI) in pathology, allowing precise TLS detection and quantification^13,14^. This integration has emphasized TLS detected or quantified by AI as a prognostic factor for overall survival across various cancer types^15–17^. However, limited research has validated AI models for assessing TLSs as predictive biomarkers in patients undergoing ICI treatment.

This study aimed to develop an AI model for objective TLS evaluation in H&E-stained WSIs and to explore its correlation with survival outcomes in NSCLC patients treated with ICIs.

## MATERIALS AND METHODS

### Development of AI-powered H&E WSI analyzer for immune phenotype and TLS analysis

The AI-powered H&E WSI analyzer, Lunit SCOPE IO (Lunit, Seoul, Republic of Korea), was developed to perform spatial analysis of the TME across more than 26 cancer types. The analyzer consists of two models: a cell detection model and a tissue segmentation model.

The cell detection model was trained on 5,609 WSIs, with 2,828,448 annotated cells, including 1,188,625 tumor cells and 706,320 lymphocytes. The tissue segmentation model was developed using 18,935 WSIs, covering a total of 1.93 × 10^10^μm², including 1.11 × 10^10^μm² of cancer area (CA), 7.95 × 10^9^ um^2^ of cancer stroma (CS), and 2.39 × 10^8^μm² corresponding to TLS regions. The model architecture is based on DeepLabV3+^18^ with a ConvNeXt-tiny^19^ backbone. To enhance generalizability across different scanners and staining variations, the model employs RandStainNA^20^, a hybrid method that integrates stain normalization and augmentation.

The cell detection model identifies tumor cells and lymphocytes, while the tissue segmentation model classifies tissue regions at pixel level into CA, CS, or TLS. By integrating data from both models, the analyzer calculates intratumoral TIL (iTIL) densities and stromal TIL (sTIL) densities based on lymphocyte counts within CA and CS. Each WSI was partitioned into 0.25mm^2^ grids.

Using TIL densities, the analyzer classified immune phenotypes (IPs) according to the following criteria: inflamed, iTIL density ≥ 130/mm2; immune-excluded, iTIL density < 130/mm2 with sTIL density ≥ 260/mm2; and immune-desert, where TIL densities fall below these thresholds in both areas. A WSI was classified as inflamed if ≥33.3% of the grids exhibited the inflamed phenotype and immune-excluded if ≥33.3% of its grids displayed the immune-excluded phenotype while the inflamed phenotype remained <33.3%. Otherwise, the WSI was categorized as immune-desert^21^. Inflamed score of WSI was defined by the number of grids annotated to inflamed phenotype divided by total analyzed grids in WSI.

The analyzer identified individual TLS clusters and quantified their total area within each WSI. TLS area was defined as the cumulative area of all TLS clusters in a given WSI. Samples were classified into two groups: TLS(+) for those with TLS, and TLS(−) for those without. Additionally, the TLS proportion (TLSP)—calculated as the ratio of TLS area to the combined area of CA and CS regions—was determined.

### Assessment of the AI-determined TLS and its association with genetic mutations and immunological characteristics in publicly available dataset

The AI-determined TLS presence was further analyzed in relation to genetic mutations and immunological characteristics using TCGA NSCLC data (*N*=924). We integrated TLS status with mutational data from cBioPortal and immune cell infiltration profiles from CIBERSORT. Additionally, to explore differences in the activity of relevant biological pathways between TLS(+) and TLS(-) samples, differential gene expression analysis and gene set enrichment analysis (GSEA) were performed. Additional details are provided in Supplementary Method 1.

### Comparative assessment of TLS presence by a pathologist and an AI Model in real-world data

For comparative assessment, we retrospectively collected H&E-stained slides and clinicopathological data from advanced-stage NSCLC patients treated at Northwestern Memorial Hospital (NMH) in Chicago, IL, USA. These patients received either ICI monotherapy or a combination of ICI and chemotherapy for palliative purposes and had pretreatment tissue biopsy slides—obtained within 547 days prior to the initiation of the nearest ICI treatment—available for scanning. All slides were scanned at 40x magnification using a Leica GT450 slide scanner. The study protocol was approved by the local Institutional Review Board (IRB) of NMH (STU00207117) and adhered to the Declaration of Helsinki.

All WSIs were reviewed by a pathologist, W. J., to confirm the quality of the WSIs as well as the presence of TLS. When evaluating the TLS status in the lymph nodes (LNs), TLSs were only considered if they were admixed with tumor cells and located distant from the residual parenchyma, ensuring the exclusion of innate lymphoid follicles. Following the assessment of the presence of TLS by a pathologist, the identical WSIs were evaluated by the AI-powered H&E WSI analyzer for TLS.

### Evaluation of TLS on clinical outcomes of ICI treatment

Differences in progression-free survival (PFS), overall survival (OS), and objective response rate (ORR) were analyzed according to TLS presence, as identified by the AI analyzer and pathologist.

The TLSP and the number of TLS clusters, quantified by the AI analyzer, were further analyzed to assess their impact on survival outcomes. The summary of the study process is shown in Figure 1A with the representative image of the TLS detection by the AI analyzer is shown in Figure 1B.

**Figure 1.**
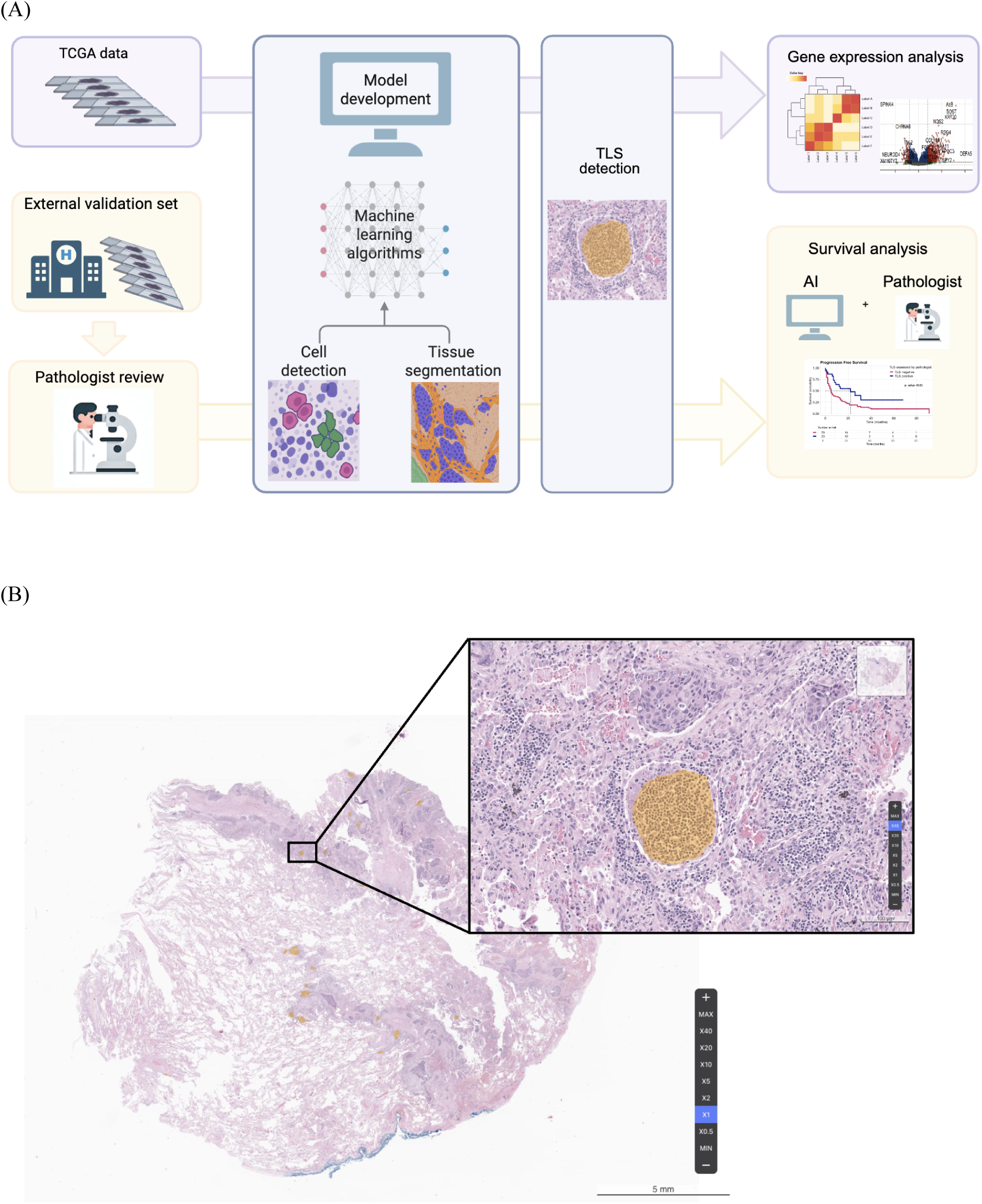
Development of an AI-powered TLS analyzer in NSCLC. (A) Schematic overview of the AI-powered TLS analyzer development and the workflow used in the current study. (B) Representative example of a TLS detection by the AI analyzer. The figure shows an image of a TLS (+) case where the AI analyzer identified TLS at low magnification (left) and high magnification (right). The TLS area detected by the AI analyzer is highlighted in orange. AI, artificial intelligence; NSCLC, non-small-cell lung cancer; TLS, tertiary lymphoid structure.

### Statistical analysis

The agreement between the pathologist and AI model interpretations was evaluated using accuracy and Cohen’s kappa. Survival outcomes were analyzed with Kaplan-Meier curves and log-rank tests, while Cox proportional hazard models were used to calculate hazard ratios (HRs) with 95% confidence intervals (CI). For categorical variables, comparisons were made using the chi-squared test or Fisher’s exact test, as appropriate. For continuous variables, differences in means or medians were assessed using the Student’s t-test or the non-parametric Mann-Whitney U test. All statistical analyses, conducted in R software, version 4.4.1 (R Foundation for Statistical Computing, Vienna, Austria), considered P < 0.05 statistically significant. Data analysis spanned August 2024 to April 2025.

## RESULTS

### Association between AI-determined TLS and genetic mutations, immune characteristics

Out of 924 TCGA-sourced NSCLC samples, an AI model identified TLS in 779 slides (84.3%). Subsequently, cBioPortal mutational data enabled the inclusion of 910 samples for further analysis. In this subset, TLS(+) was 85.3% (776/910). The proportions of LUAD were 49.4% (383/776) and 55.2% (74/134) for TLS (+) and TLS (-) respectively. The occurrence of mutations were as follows: EGFR (55 cases), KRAS (148 cases), BRAF (36 cases), ERBB2 (4 cases), and MET splicing variants (6 cases). Additionally, translocations were identified in ALK (4 cases), ROS1 (6 cases), RET (2 cases), and NTRK2 (1 case), with no NTRK1/3 translocations detected. None of these mutational patterns were found to be significantly different between the TLS groups (Supplementary Table 1).

Using CIBERSORT, a total of 903 cases were included in this analysis. The TLS (+) group demonstrated a significant positive correlation with memory B cells (fold change [fc] 1.78, *p*=0.005), CD8 T cells (fc 1.34, *p*=0.001), naive B cells (fc 1.28, *p*=0.008), regulatory T cells (fc 1.27, *p*=0.012), and follicular helper T cells (fc 1.15, *p*=0.001), while a negative correlation was observed with Monocytes (fc 0.725, *p*=0.024) and neutrophils (fc 0.595, *p*=0.001) when compared to the TLS (-) group (Figure 2A).

**Figure 2.**
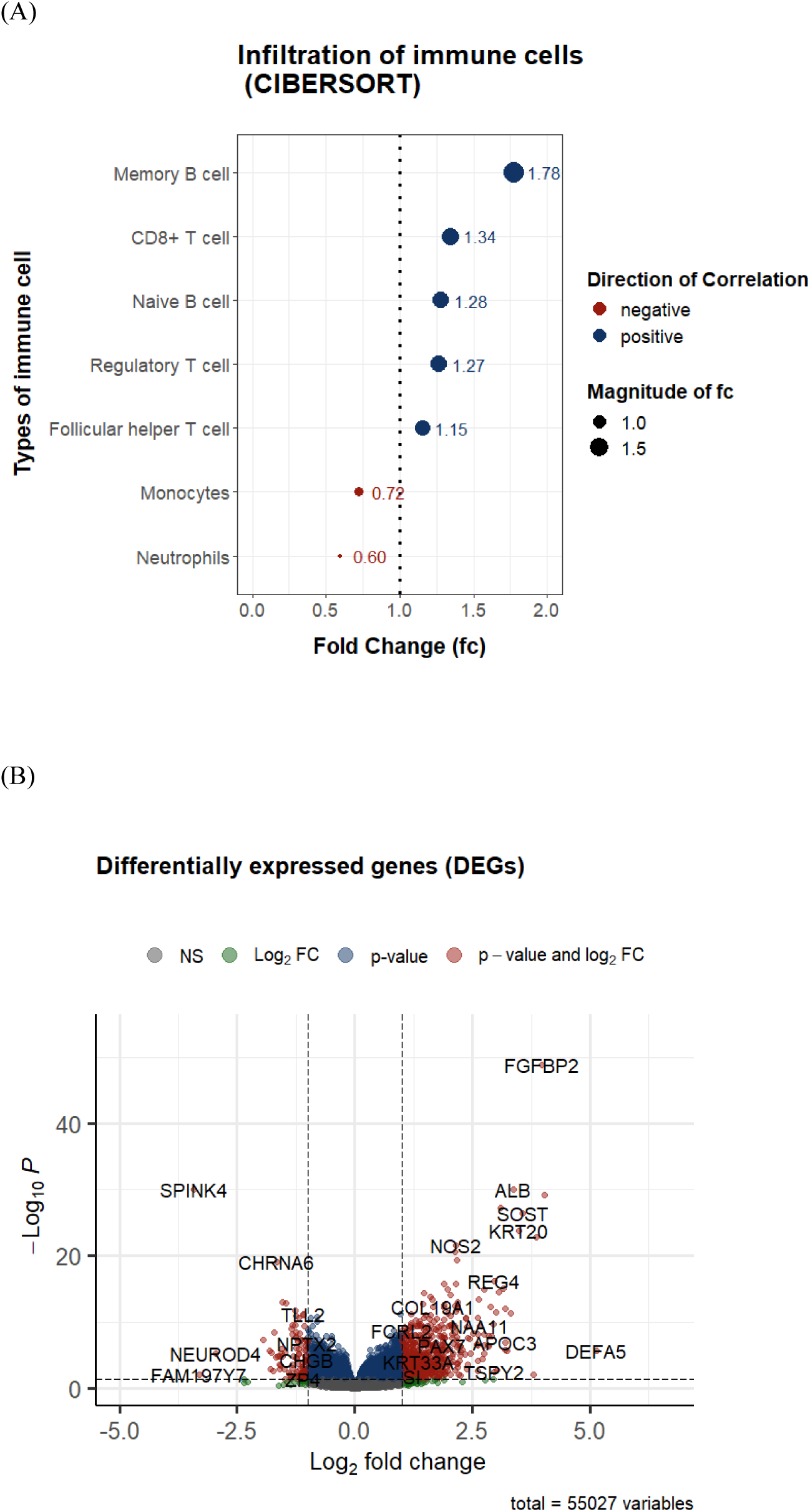
(A) Correlation between AI-determined TLS presence and infiltration of immune cells in non-small cell lung cancer (NSCLC) samples from The Cancer Genome Atlas (TCGA) dataset. (B) Volcano plot of differentially expressed genes identified between the tertiary lymphoid structure (TLS) (+) and TLS (-) group.

From the results of differential gene expression analysis, a total 868 genes that differentially expressed between the TLS (+) and TLS (-) groups were found with cutoff of p-value < 0.05 and fc > 1.0 (Figure 2B). Subsequent GSEA analysis of Hallmark pathways or biological process components of gene ontology (C5 GO:BP) gene sets showed TLS (+) group was positively associated with immunoglobulin production (normalized enrichment score [NES] 2.17, *p_adj_*=0.02) and B cell receptor signaling pathway (NES 1.85, *p_adj_*=0.02). Pathways displaying the top 10 absolute values of NES with *p_adj_* < 0.05 are summarized in Supplementary Table 2.

### Analysis of AI vs pathologist - determined TLS in patients of NSCLC

A total of 102 patients were included in the study, while 60 cases were excluded for the following reasons: 18 cases were obtained more than 1.5 years (547 days) before ICI treatment initiation; 18 cases contained only LNs; 16 cases received immunotherapy with curative intent; six cases were post-treatment biopsy samples; and two were lost to follow-up (Supplementary Figure 1). Baseline clinicopathologic characteristics based on TLS assessment by the AI analyzer are summarized in Table 1, while those based on TLS assessment by a pathologist are detailed in Supplementary Table 3. 30.3% (31/102) and 22.5% (23/102) of cases were found to contain TLS (TLS(+)) as determined by the AI analyzer and a pathologist respectively. The AI model achieved an accuracy of 88.2% (95% confidence interval [CI], 80.4%–93.8%), with a sensitivity of 0.87 and specificity of 0.91 in identifying the presence of TLS, compared to the pathologist’s interpretation (Figure 3A). The concordance between the pathologist’s assessment and the AI model, as evaluated by Cohen’s kappa, was 0.70. Among the ten false-positive cases identified by the AI analyzer, the analyzer correctly detected lymphocytes in some cases; however, they did not form definite aggregates (Supplementary Figure 2).

**Figure 3.**
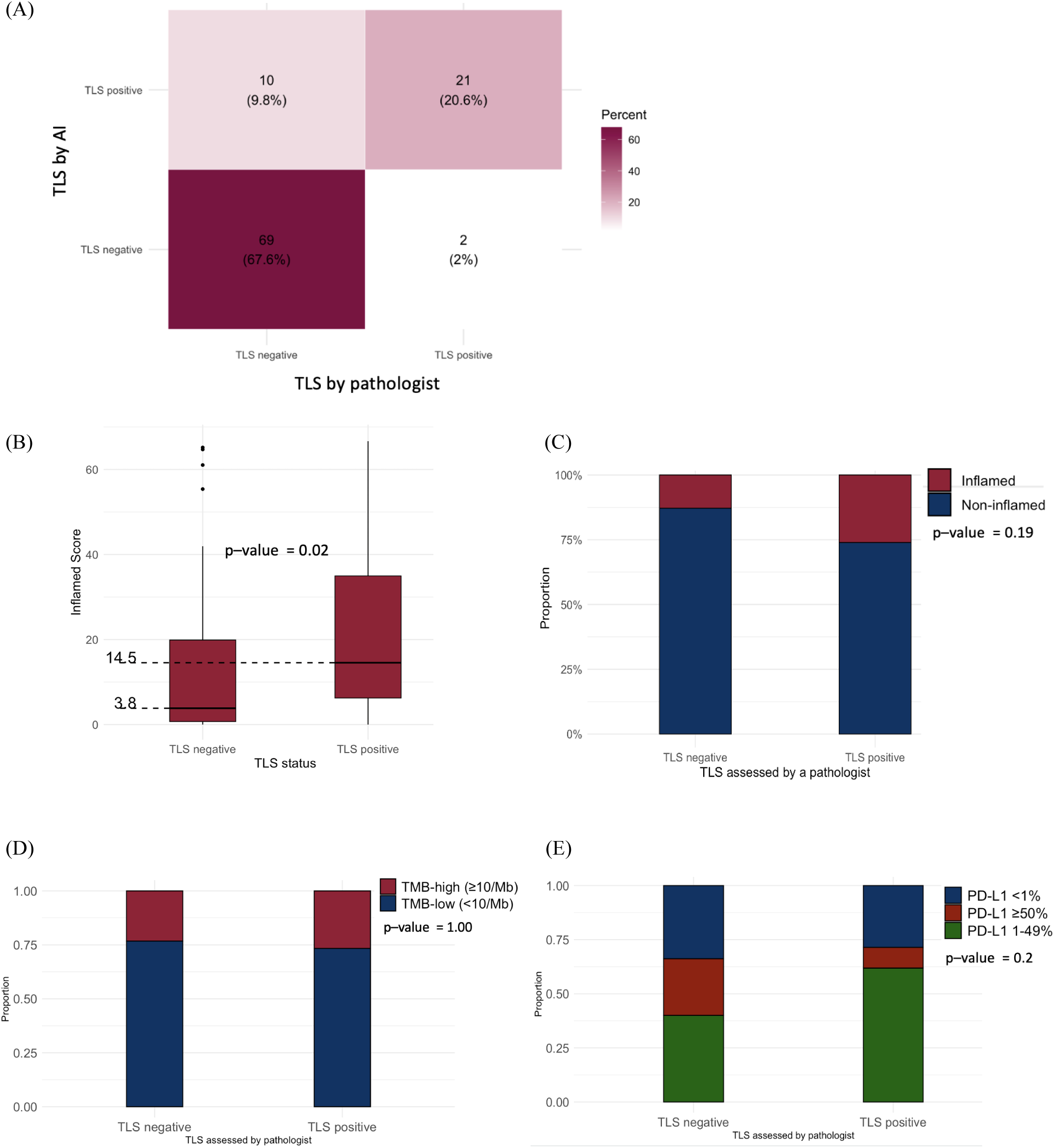
Analysis of AI- and pathologist-determined TLS in NSCLC patients. (A) Confusion matrix comparing TLS presence detection between the AI analyzer and pathologist interpretation. (B) Distribution of inflamed scores (IS) according to TLS presence as assessed by the pathologist. Thick horizontal lines represent median IS values. (C) Immune phenotypes, (D) TMB levels, and (E) proportion of PD-L1 expression levels according to TLS presence assessed by the pathologist. AI, artificial intelligence; NSCLC, non-small-cell lung cancer; PD-L1, programmed cell death ligand 1; TLS, tertiary lymphoid structure; TMB, tumor mutational burden.

**Table 1.**
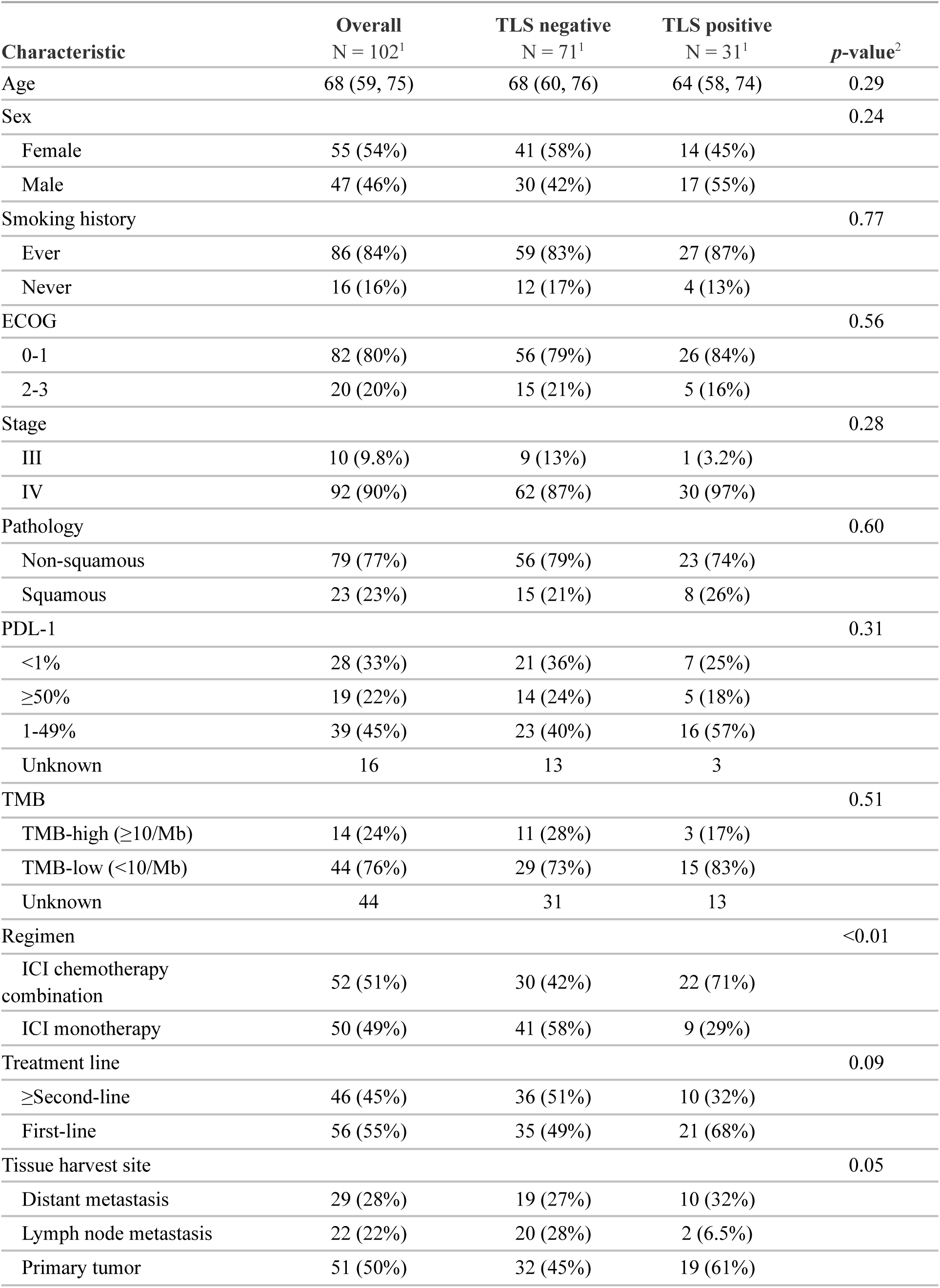
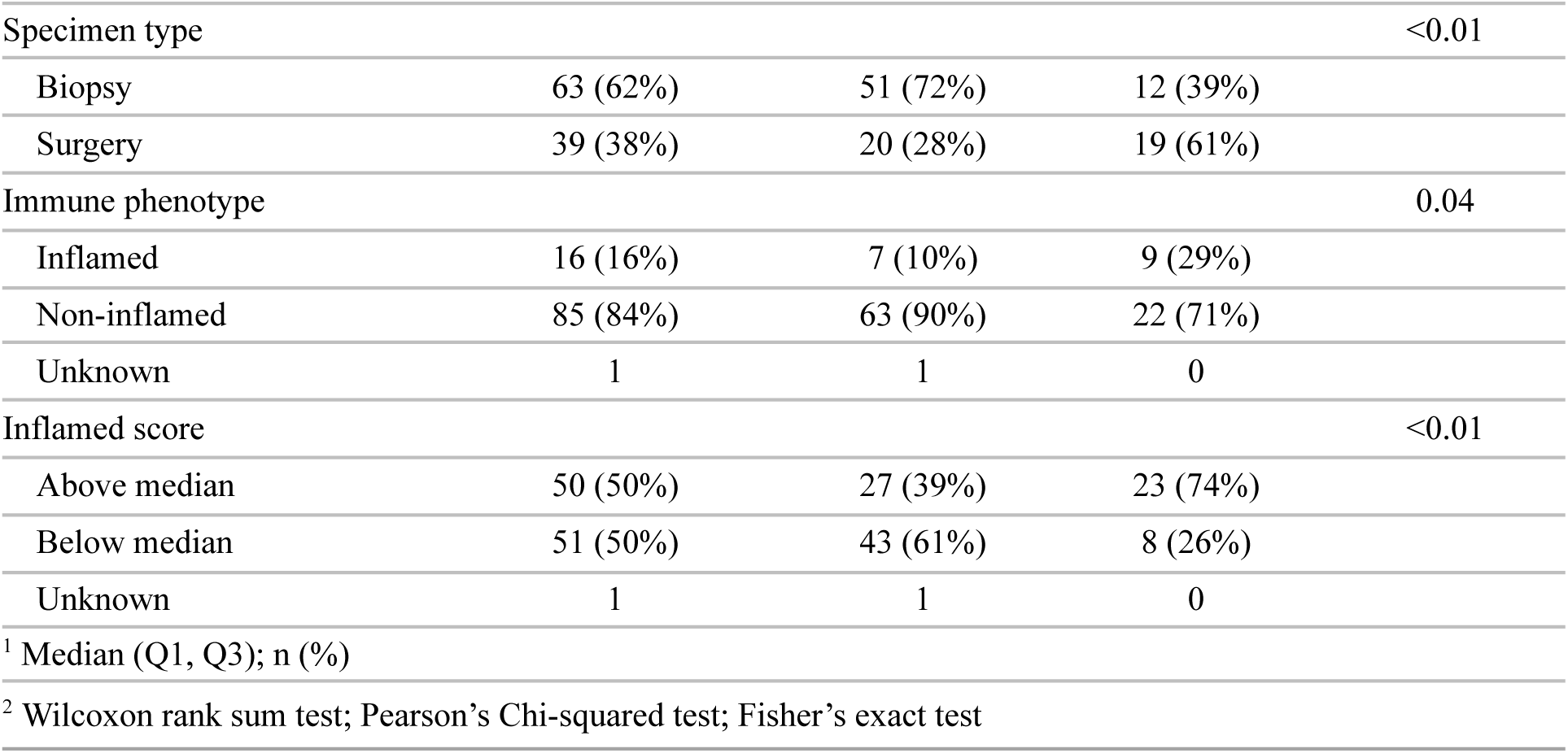
Patient characteristics with TLS assessed by AI analyzer. AI, artificial intelligence; TLS, tertiary lymphoid structure.

### The distribution of inflamed scores, immune phenotypes, TMB levels, and PD-L1 expression levels between the TLS (+) and TLS (-) determined by a pathologist

The TLS (+) group showed a higher median inflamed score (14.5, interquartile range [IQR] 6.25 - 34.97) compared to the TLS (-) group (3.8, IQR 0.63 - 19.88, *p*=0.02). The TLS (+) group had a higher proportion of the inflamed immune phenotype (26% vs. 13%). There was no significant difference in the categorical distribution of TMB and PD-L1 expression level between the two groups (Table 1, Figure 3 B-E).

### Analysis of ICI treatment outcomes based on AI- and pathologist-determined TLS presence

The median follow-up duration was 40.0 months (mo). TLS (+) status was associated with significantly favorable survival outcomes for both PFS (by pathologist: HR 0.45, 95% CI 0.25–0.82, *p* < 0.01, mPFS 22 vs 5 mo; by AI: HR 0.49, 95% CI 0.29–0.82, *p* < 0.01, mPFS 21 vs 5 mo) and OS (by pathologist: HR 0.43, 95% CI 0.23–0.78, *p* <0.01, mOS 37 vs 13 mo; by AI: HR 0.55, 95% CI 0.33–0.92, *p*=0.02, mOS 26 vs 13 mo) compared to the TLS (-) group. There was no significant difference in ORR between the TLS (+) and TLS (-) groups, as determined by the pathologist (17.4% vs. 17.7%) and the AI analyzer (19.4% vs. 16.9%) (Figure 4 A and B).

**Figure 4.**
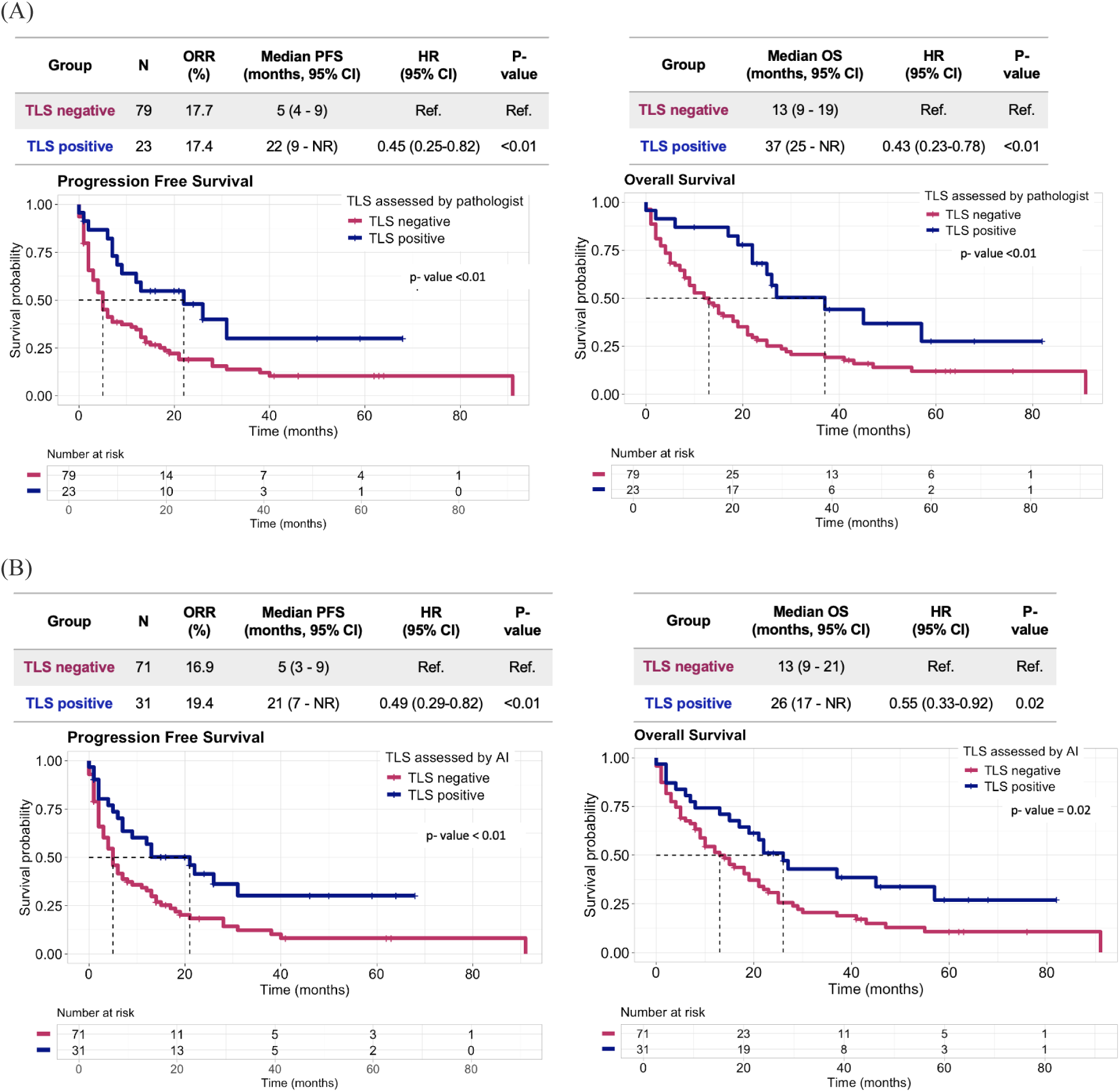
Kaplan-Meier survival analysis of PFS (left) and OS (right) after ICI treatment, according to the presence of TLS assessed by (A) a pathologist and (B) AI analyzer. P-values were calculated using a two-sided log-rank test. The Cox proportional hazards model was used for calculation of HRs and corresponding 95% CIs. AI, artificial intelligence; CI, confidence interval; HR, hazard ratio; ICI, immune checkpoint inhibitor; NR, not reached; ORR, objective response rate; OS, overall survival; PD-L1, programmed death ligand-1; PFS, progression-free survival; TLS, tertiary lymphoid structure.

When comparing patients classified as TLS (+) by both a pathologist and the AI analyzer with those classified as TLS (–) by both, the TLS (+) group had significantly better PFS (HR 0.39, 95% CI 0.20–0.74, *p* < 0.01) and OS (HR 0.42, 95% CI 0.22–0.78, *p* < 0.01) (Supplementary Figure 3).

Both pathologist- and AI-determined TLS (+) status remained statistically associated with longer PFS and OS after adjusting for clinicopathologic characteristics. TMB was excluded from the multivariate Cox regression analysis due to a high proportion of missing data (44 of 102 cases, 43.1%). Detailed results of the multiple univariate and multivariable analyses are presented in Supplementary Table 4 and 5, respectively.

### Survival analysis based on the presence of TLS across the treatment regimen type (ICI monotherapy vs. ICI combined with chemotherapy), compared to the performance of PD-L1 expression levels

For the 50 patients who underwent ICI monotherapy, TLS (+) status was associated with significantly favorable survival outcomes for both PFS (by pathologist: HR 0, 95% CI 0-inf, *p* < 0.01; by AI: HR 0.41, 95% CI 0.17–1.15, *p* = 0.08) and OS (by pathologist: HR 0, 95% CI 0–inf, *p* < 0.01; by AI: HR 0.54, 95% CI 0.21–1.38, *p* = 0.19) compared to the TLS (-) group (Figure 5A, Supplementary Figure 4A). The PD-L1 high (≥50%) group did not show a significant difference in PFS and OS compared to the PD-L1 low (<50%) group (Figure 5B).

**Figure 5.**
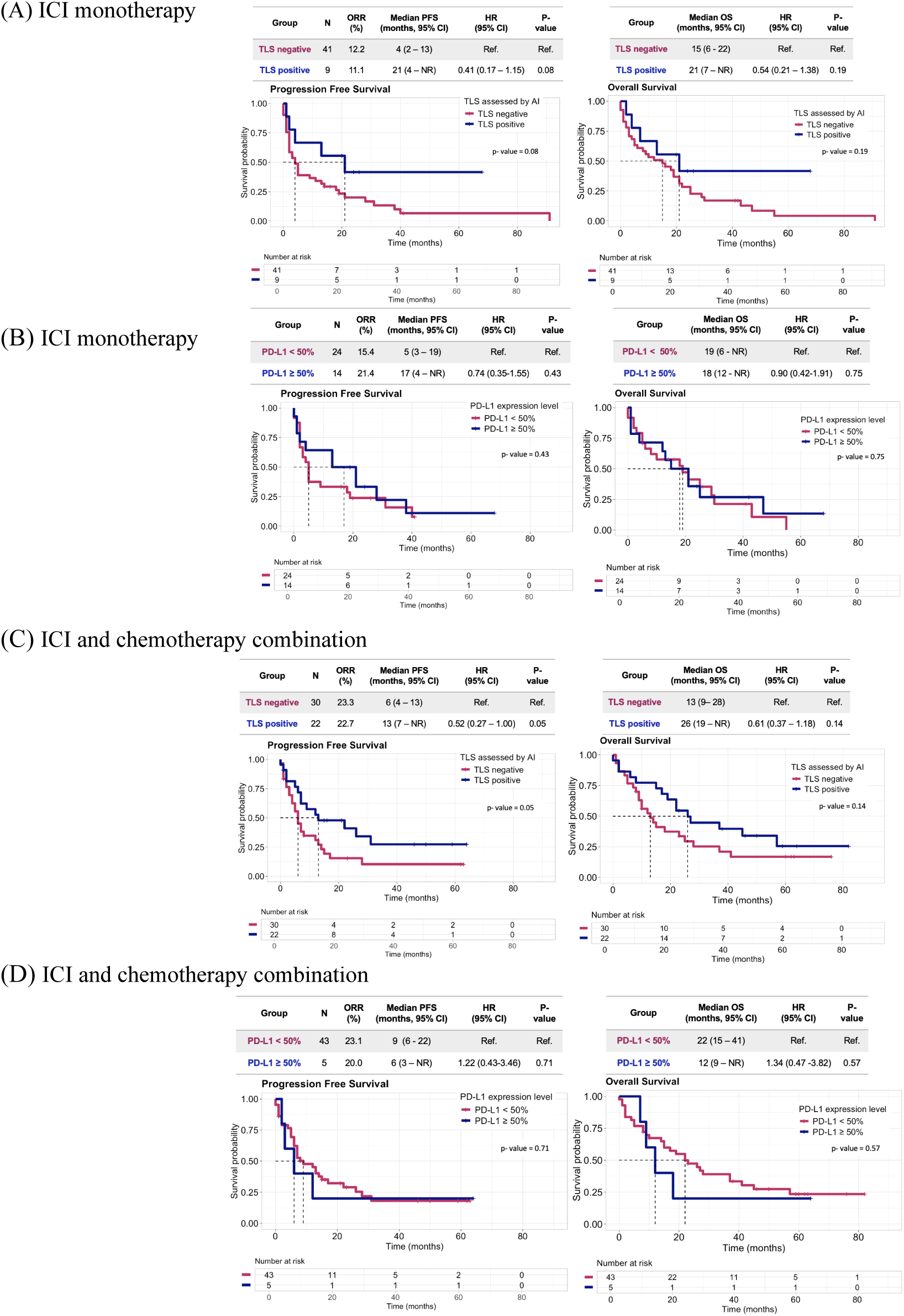
Kaplan-Meier survival analysis of PFS (left) and OS (right) following ICI monotherapy based on (A) the presence of TLS assessed by AI analyzer and (B) PD-L1 expression level. Kaplan-Meier survival analysis of PFS (left) and OS (right) following ICI and chemotherapy combination based on (C) the presence of TLS assessed by a AI analyzer and (D) PD-L1 expression level. P-values were calculated using a two-sided log-rank test. The Cox proportional hazards model was used for calculation of HRs and corresponding 95% CIs. AI, artificial intelligence; CI, confidence interval; HR, hazard ratio; ICI, immune checkpoint inhibitor; NR, not reached; ORR, objective response rate; OS, overall survival; PD-L1, programmed death ligand-1; PFS, progression-free survival; TLS, tertiary lymphoid structure.

For the 52 patients treated with ICI combined with chemotherapy, TLS (+) status was associated with numerically favorable PFS (by pathologist: HR 0.65, 95% CI 0.33-1.26, *p* = 0.21, mPFS 13 vs 6 mo; by AI analyzer: HR 0.52, 95% CI 0.27–1.00, *p* = 0.05, mPFS 13 vs 6 mo) and OS (by pathologist: HR 0.64, 95% CI 0.33–1.26, *p* = 0.19, mOS 26 vs 13 mo; by AI analyzer: HR 0.61, 95% CI 0.37–1.18, *p* = 0.14, mOS 26 vs 13 mo) compared to the TLS (-) group (Figure 5C, Supplementary Figure 4B). The PD-L1 high (≥50%) group did not show a significant difference in PFS and OS compared to the PD-L1 low (<50%) group (Figure 5D).

### Survival analysis based on the presence of TLS across the lines of treatment (first-line vs second-line and beyond)

Among 56 patients who received immunotherapy as a first-line treatment, TLS (+) status was associated with numerically better PFS (by pathologist: HR 0.60, 95% CI 0.29–1.22, *p* = 0.16, mPFS 22 vs 11 mo; by AI analyzer: HR 0.62, 95% CI 0.33–1.20, *p* = 0.15, mPFS 21 vs 8 mo) and OS (by pathologist: HR 0.61, 95% CI 0.30–1.24, *p* = 0.17, mOS 37 vs 18 mo; by AI analyzer: HR 0.76, 95% CI 0.39–1.45, *p* = 0.40, mOS 26 vs 18 mo) (Supplementary Figure 5 A and B). Among 46 patients who received immunotherapy as second-line or beyond treatment, TLS(+) status was associated with significantly better PFS (by pathologist: HR 0.31, 95% CI 0.09–1.03, *p* = 0.04; by AI analyzer: HR 0.41, 95% CI 0.17–0.99, *p* = 0.05) and OS (by pathologist: HR 0.26, 95% CI 0.08–0.85, *p* = 0.02; by AI analyzer: HR 0.43, 95% CI 0.18–1.03, *p* = 0.05) (Supplementary Figure 5 C and D).

### Survival analysis based on the presence of TLS across the PD-L1 expression subgroups (PD-L1 positive ( ≥1%) vs PD-L1 negative (<1%))

Among 58 patients with positive PD-L1 expression (≥1%), TLS (+) status was associated with significantly better PFS (by pathologist: HR 0.48, 95% CI 0.23–0.98, *p* = 0.05; by AI analyzer: HR 0.43, 95% CI 0.22–0.83, *p* < 0.01) and OS (by pathologist: HR 0.34, 95% CI 0.15–0.77, *p* < 0.01; by AI analyzer: HR 0.40, 95% CI 0.20–0.80, *p* < 0.01) (Supplementary Figure 6 A and B). Among 28 patients with negative PD-L1 expression ( <1%), TLS (+) status was associated with numerically better PFS (by pathologist: HR 0.37, 95% CI 0.11–1.26, *p* = 0.09, mPFS 31 vs 6 mo; by AI analyzer: HR 0.50, 95% CI 0.17–1.51, *p* = 0.21, mPFS 22 vs 6 mo) and no significant difference in OS (Supplementary Figure 6 C and D).

### Survival analysis based on the presence of TLS by tissue harvest site (primary vs metastatic lesion)

Among the 51 patients whose tissue was harvested from the primary lesion, TLS (+) status showed numerically longer PFS (by pathologist: HR 0.72, 95% CI 0.35–1.47, *p* = 0.38, mPFS 13 vs 5 mo; by AI analyzer:; HR 0.56, 95% CI 0.28–1.09, *p* = 0.08, mPFS 13 vs 5 mo) and OS (by pathologist: HR 0.69, 95% CI 0.33–1.43, *p* = 0.31, mPFS 25 vs 15 mo; by AI analyzer: HR 0.74, 95% CI 0.38–1.45, *p* = 0.37, mPFS 22 vs 18 mo) compared to the TLS (-) group (Figure 6 A and B).

**Figure 6.**
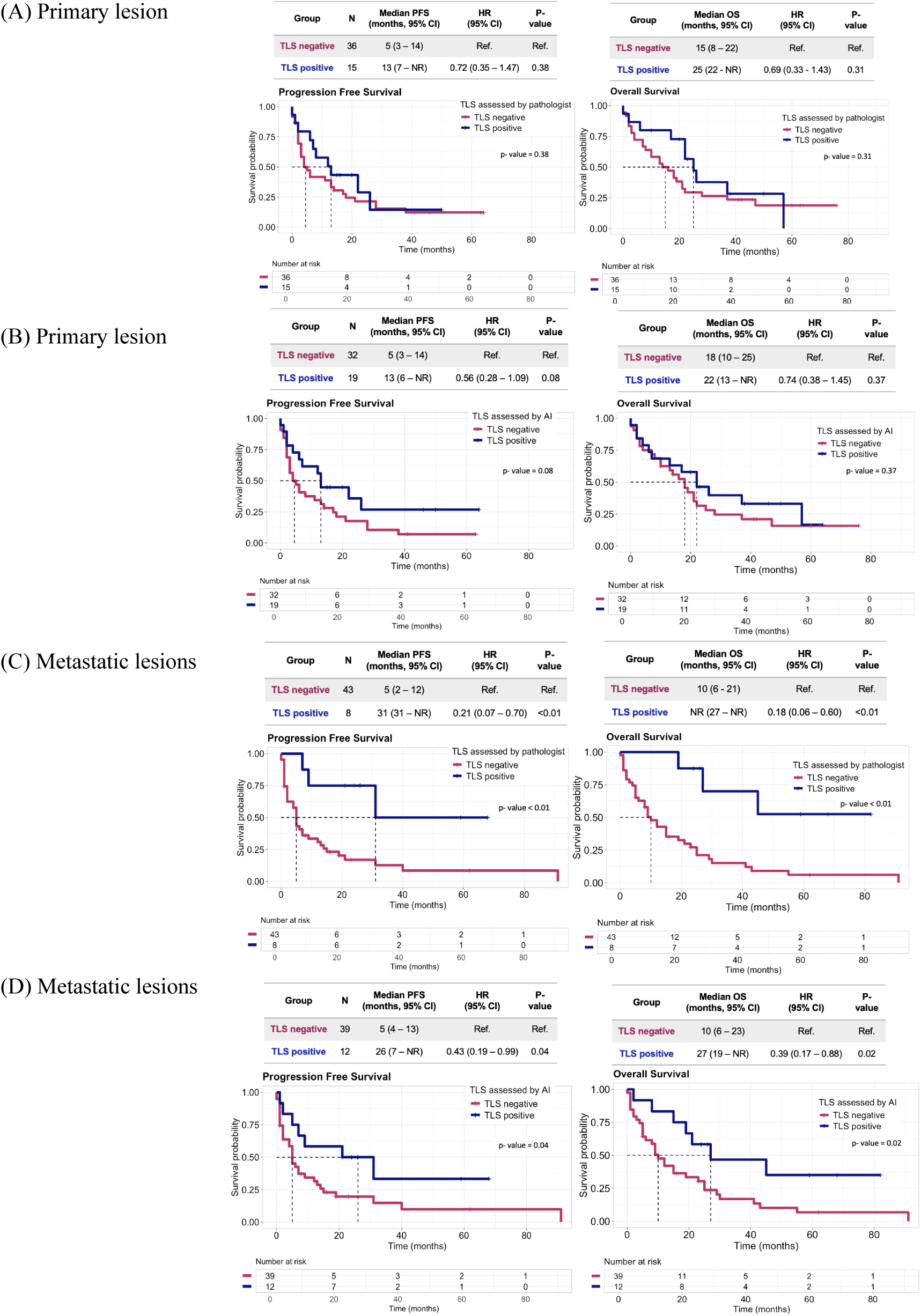
Kaplan-Meier survival analyses of PFS (left) and OS (right) following ICI treatment in patients with tissue harvested from the primary lesion, stratified by the presence of TLS as assessed by (A) a pathologist and (B) an AI analyzer. Kaplan-Meier survival analyses of PFS (left) and OS (right) following ICI treatment in patients with tissue harvested from metastatic lesions, stratified by the presence of TLS as assessed by (C) a pathologist and (D) an AI analyzer. P-values were calculated using a two-sided log-rank test. The Cox proportional hazards model was used for calculation of HRs and corresponding 95% CIs. AI, artificial intelligence; CI, confidence interval; HR, hazard ratio; ICI, immune checkpoint inhibitor; NR, not reached; ORR, objective response rate; OS, overall survival; PD-L1, programmed death ligand-1; PFS, progression-free survival; TLS, tertiary lymphoid structure.

For the other 51 patients whose tissue was harvested from metastatic lesions, TLS (+) status was associated with significantly longer PFS (by pathologist: HR 0.21, 95% CI 0.07–0.70, *p* <0.01; by AI analyzer: HR 0.43, 95% CI 0.19–0.99, *p* = 0.04) and OS (by pathologist: HR 0.18, 95% CI 0.06–0.60, *p* <0.01; by AI analyzer: HR 0.39, 95% CI 0.17–0.88, *p* = 0.02) compared to the TLS (-) group (Figure 6 C and D).

### Survival analysis based on TLS size and number of clusters (above median vs below median)

In the TLS (+) group, the median total TLS size was 0.12 mm² (IQR: 0.06–0.31 mm²). The TLSP was also calculated, yielding a median of 0.54% (IQR: 0.21 – 1.56%). The median number of TLS clusters was 2 (IQR: 1–5). Among the TLS (+) group, there was no significant difference in PFS or OS between patients with TLSP above or below the median (Supplementary Figure 7 A). Additionally, there was no significant difference in PFS or OS between patients whose number of TLS clusters was above or below the median (Supplementary Figure 7 B).

### Survival analysis based on immune score (above median vs below median) and immune phenotype (inflamed vs non-inflamed)

Among 101 patients with available immune scores and immune phenotypes, immune score above median was associated with numerically better PFS (HR 0.65, 95% CI 0.42–1.02, *p* = 0.06, mPFS 12 vs 5) and OS (HR 0.72, 95% CI 0.46–1.13, *p* = 0.15, mPFS 19 vs 13) (Supplementary Figure 8 A). Inflamed immune phenotype was associated with numerically better PFS (HR 0.56, 95% CI 0.29–1.06, *p* = 0.07, mPFS 26 vs 6) and OS (HR 0.63, 95% CI 0.34–1.18, *p* = 0.15, mPFS 28 vs 14) (Supplementary Figure 8 B).

## DISCUSSION

The presence of TLS, as detected by our AI-based analyzer, was validated by the enrichment of specific types of tumor-infiltrating immune cells in TLS (+) samples, including memory B cells, CD8⁺ T cells, naive B cells, regulatory T cells, and follicular helper T cells. Furthermore, the GSEA revealed an association of the TLS (+) group with immunoglobulin production, and the B cell receptor signaling pathway. This may suggest active cooperation between these cell subsets in generating an effective anti-tumor immune response^11,22,23^. This finding aligns with previous research reporting that the presence of the intratumoral B cell, plasma cell, PD-1 high dysfunctional CD8+ T cells that are associated with the presence of TLSs was shown to predict response to ICIs in NSCLC^24–27^.

To the best of our knowledge, we developed the first AI model for detecting the presence of TLS in WSI, with clinical validation from real-world data confirming its predictive value for survival outcomes in advanced-stage NSCLC patients receiving immunotherapy. Notably, we are the first to report that the predictive performance of TLS was enhanced when assessed from WSI of metastatic lesions.

Several deep learning (DL) models have been developed to automate TLS segmentation from H&E slides across various tumor types, including lung cancer^28^. And several studies have validated the association between these automated assessments and clinical responses to immunotherapy. For example, Chen et al. presented a DL model for automated TLS segmentation using a dataset of matched multiplex IHC and H&E WSI^29^. They demonstrated that NSCLC patients who underwent neoadjuvant therapy (combining anti-PD-1 and chemotherapy or anti-PD-1 and apatinib) and achieved a significant tumor reduction (>90%) in pathological response had a markedly higher TLS ratio—defined as the segmented TLS area divided by total tissue area—than those who did not respond similarly. In a separate study, Rakekee et al. introduced an AI-based method for quantifying TLSs in digitized H&E slides. Among NSCLC patients treated with first-line or subsequent lines of anti-PD-(L)1 monotherapy, those with ≥0.01 TLS per mm² demonstrated significantly higher ORR, as well as longer PFS and OS. However, these findings were reported only as an abstract^30^.

Compared to previous studies, our research incorporates clinical validation using real-world data from advanced-stage NSCLC patients treated with either ICI monotherapy or ICI combined with chemotherapy, across all lines of therapy, complemented by subgroup analyses. The presence of TLS, rather than its density, was consistently associated with favorable survival outcomes, regardless of PD-L1 status, treatment regimen (ICI monotherapy vs. combination therapy), lines of therapy (first-line vs. subsequent lines), or tissue harvest site (primary vs. metastatic). These findings raise the possibility that TLS presence could serve as a universal and independent biomarker for predicting immunotherapy response in advanced NSCLC. This is particularly relevant as currently approved biomarkers—largely validated in the frontline setting for ICI monotherapy—often lose predictive value beyond frontline therapy or in combination regimens^31–33^, underscoring the importance of identifying additional predictive markers.

Interestingly, TLS demonstrated superior performance in stratifying survival outcomes for both PFS and OS when detected in tissue samples from metastatic lesions. While studies on TLSs in metastatic sites remain limited, they offer valuable insights, reinforcing the concept that antitumor immunity remains active even at the metastatic stage^8^. TLS density in metastatic sites is known to vary widely and often parallels that of the primary tumor^9^. However, emerging research suggests that TLS maturation may differ depending on the metastatic site. For example, in microdissected follicles from melanoma skin metastases, B lymphocytes exhibited local antigen-driven responses with isotype switching—not only to IgG subclasses but also frequently to IgA—despite the non-mucosal location of the metastasis^34^. This phenomenon, also observed in TLS-rich primary NSCLC tumors, may be driven by elevated levels of TGFβ^35^, a known switch factor for IgA. To our knowledge, this is the first study to report the predictive value of TLS presence in metastatic lesions of NSCLC. Further research is warranted to elucidate the biology of TLS maturation in metastatic sites and its implications for predicting response to immunotherapy.

This study has limitations. First, being a retrospective study conducted at a single hospital. Second, the AI model’s accuracy was determined by comparing results from only one pathologist. Given the interobserver variability of TLS with H&E slides, results from a single pathologist could potentially be inaccurate. Multiple pathologists’ evaluations would have added an additional layer of validation. Third, the clinical validation used samples from various sites, from primary tumors and LNs to distant metastatic sites such as the brain, liver, or bone. Although the effect of TLS on survival outcomes according to biopsy sites has not been investigated in NSCLC patients, the potential effect of TLS on survival outcomes may differ according to biopsy sites, as reported in other cancer types such as breast cancer, colorectal cancer, or renal cell carcinoma.

Despite these limitations, our study contributes to the understanding of TLS in the context of NSCLC and its potential as a biomarker for ICI treatment response. However, it also highlights the need for further research to overcome the current limitations and to explore the implications of TLS in NSCLC treatment more comprehensively.

## Supporting information

Supplementray_TLS

## Data Availability

All data produced in the present study are available upon reasonable request to the authors.

## Declarations

### Ethics approval and consent to participate

For comparative assessment, we retrospectively collected hematoxylin and eosin (H&E)-stained slides and clinicopathological data from patients with advanced-stage non-small cell lung cancer (NSCLC) treated at Northwestern Memorial Hospital (NMH) in Chicago, IL, USA. Eligible patients received immune checkpoint inhibitor (ICI) monotherapy or a combination of ICI and chemotherapy for palliative intent and had pretreatment tissue biopsy slides—acquired within 547 days prior to initiation of the nearest ICI regimen—available for scanning. All slides were digitized at 40× magnification using a Leica GT450 slide scanner. The study protocol was reviewed and approved by the Institutional Review Board (IRB) of NMH (STU00207117) and was conducted in accordance with the Declaration of Helsinki. Given the retrospective nature of the study, the need for informed consent for participation and publication was waived.

### Availability of data and materials

The datasets generated and/or analyzed during the current study are not publicly available due to institutional data-sharing policies but are available from the corresponding author on reasonable request and subject to IRB and institutional approval.

### Authors’ contributions

Leeseul Kim: Conceptualization, Ideas, Methodology, Software, Validation, Formal Analysis, Investigation, Resources, Data Curation, Writing – Original Draft, Visualization, Project Administration. Sangwon Shin: Conceptualization, Ideas, Methodology, Software, Validation, Formal Analysis, Investigation, Resources, Writing – Review & Editing, Visualization. Behtash G. Nezami: Resources. Borislav Alexiev: Resources. Soo Ick Cho: Supervision, Project Administration. Sanghoon Song: Software, Formal Analysis. Wonkyung Jung: Methodology, Resources. Lee AD Cooper: Resources. Donghoon Shin: Data Curation. Junho Song: Data Curation. Taekyu Um: Data Curation. Liam Il-Young Chung: Resources, Project Administration. Young Kwang Chae: Conceptualization, Ideas, Methodology, Supervision.

## Acknowledgements

The authors gratefully acknowledge the support of Lunit and the invaluable assistance of Dr. Jeeyun Ryu.

Portions of this work were presented in abstract form at the 2022 American Society of Clinical Oncology (ASCO) Annual Meeting (June 3–7, 2022, Chicago, USA), the IASLC 2023 World Conference on Lung Cancer (September 9–12, 2023, Singapore), the 2024 ASCO Annual Meeting (June 3–7, 2024, Chicago, USA), and the 2025 American Association for Cancer Research (AACR) Annual Meeting (April 25–30, 2025).

## Abbreviations

AI: artificial intelligence
ASCO: American Society of Clinical Oncology
BP: biological process
CA: cancer area
CIBERSORT: Cell-type Identification by Estimating Relative Subsets of RNA Transcripts
CI: confidence interval
CS: cancer stroma
DL: deep learning
EGFR: epidermal growth factor receptor
GSEA: gene set enrichment analysis
H&E: hematoxylin and eosin
HR: hazard ratio
ICI: immune checkpoint inhibitor
IHC: immunohistochemistry
iTIL: intratumoral tumor-infiltrating lymphocyte
IQR: interquartile range
LN: lymph node
LUAD: lung adenocarcinoma
mOS: median overall survival
mPFS: median progression-free survival
NES: normalized enrichment score
NMH: Northwestern Memorial Hospital
NSCLC: non-small cell lung cancer
ORR: objective response rate
OS: overall survival
PD-1: programmed cell death protein 1
PD-L1: programmed death-ligand 1
PFS: progression-free survival
RNA: ribonucleic acid
sTIL: stromal tumor-infiltrating lymphocyte
TGFβ: transforming growth factor beta
TIL: tumor-infiltrating lymphocyte
TLS: tertiary lymphoid structure
TLSP: TLS proportion
TMB: tumor mutational burden
TME: tumor microenvironment
WSI: whole slide image.

## REFERENCE

1. Liu L, Bai H, Wang C, et al. Efficacy and Safety of First-Line Immunotherapy Combinations for Advanced NSCLC: A Systematic Review and Network Meta-Analysis. Journal of Thoracic Oncology. 2021;16(7):1099–1117. doi:10.1016/j.jtho.2021.03.016

2. Arbour KC, Riely GJ. Systemic Therapy for Locally Advanced and Metastatic Non–Small Cell Lung Cancer: A Review. JAMA. 2019;322(8):764. doi:10.1001/jama.2019.11058

3. Hellmann MD, Paz-Ares L, Bernabe Caro R, et al. Nivolumab plus Ipilimumab in Advanced Non–Small-Cell Lung Cancer. N Engl J Med. 2019;381(21):2020–2031. doi:10.1056/NEJMoa1910231

4. Gogishvili M, Melkadze T, Makharadze T, et al. Cemiplimab plus chemotherapy versus chemotherapy alone in non-small cell lung cancer: a randomized, controlled, double-blind phase 3 trial. Nat Med. 2022;28(11):2374–2380. doi:10.1038/s41591-022-01977-y

5. Grossman JE, Vasudevan D, Joyce CE, Hildago M. Is PD-L1 a consistent biomarker for anti-PD-1 therapy? The model of balstilimab in a virally-driven tumor. Oncogene. 2021;40(8):1393–1395. doi:10.1038/s41388-020-01611-6

6. McGrail DJ, Pilié PG, Rashid NU, et al. High tumor mutation burden fails to predict immune checkpoint blockade response across all cancer types. Annals of Oncology. 2021;32(5):661–672. doi:10.1016/j.annonc.2021.02.006

7. Mino-Kenudson M, Schalper K, Cooper W, et al. Predictive Biomarkers for Immunotherapy in Lung Cancer: Perspective From the International Association for the Study of Lung Cancer Pathology Committee. Journal of Thoracic Oncology. 2022;17(12):1335–1354. doi:10.1016/j.jtho.2022.09.109

8. Sautès-Fridman C, Petitprez F, Calderaro J, Fridman WH. Tertiary lymphoid structures in the era of cancer immunotherapy. Nat Rev Cancer. 2019;19(6):307–325. doi:10.1038/s41568-019-0144-6

9. Dieu-Nosjean MC, Goc J, Giraldo NA, Sautès-Fridman C, Fridman WH. Tertiary lymphoid structures in cancer and beyond. Trends in Immunology. 2014;35(11):571–580. doi:10.1016/j.it.2014.09.006

10. Zhang Q, Wu S. Tertiary lymphoid structures are critical for cancer prognosis and therapeutic response. Front Immunol. 2023;13:1063711. doi:10.3389/fimmu.2022.1063711

11. Helmink BA, Reddy SM, Gao J, et al. B cells and tertiary lymphoid structures promote immunotherapy response. Nature. 2020;577(7791):549–555. doi:10.1038/s41586-019-1922-8

12. Buisseret L, Desmedt C, Garaud S, et al. Reliability of tumor-infiltrating lymphocyte and tertiary lymphoid structure assessment in human breast cancer. Modern Pathology. 2017;30(9):1204–1212. doi:10.1038/modpathol.2017.43

13. De Bree R, Meerkerk CDA, Halmos GB, et al. Measurement of Sarcopenia in Head and Neck Cancer Patients and Its Association With Frailty. Front Oncol. 2022;12:884988. doi:10.3389/fonc.2022.884988

14. Bera K, Schalper KA, Rimm DL, Velcheti V, Madabhushi A. Artificial intelligence in digital pathology — new tools for diagnosis and precision oncology. Nat Rev Clin Oncol. 2019;16(11):703–715. doi:10.1038/s41571-019-0252-y

15. Wang Y, Lin H, Yao N, et al. Computerized tertiary lymphoid structures density on H&E-images is a prognostic biomarker in resectable lung adenocarcinoma. iScience. 2023;26(9):107635. doi:10.1016/j.isci.2023.107635

16. Li Z, Jiang Y, Li B, et al. Development and Validation of a Machine Learning Model for Detection and Classification of Tertiary Lymphoid Structures in Gastrointestinal Cancers. JAMA Netw Open. 2023;6(1):e2252553. doi:10.1001/jamanetworkopen.2022.52553

17. Van Rijthoven M, Obahor S, Pagliarulo F, et al. Multi-resolution deep learning characterizes tertiary lymphoid structures and their prognostic relevance in solid tumors. Commun Med. 2024;4(1):5. doi:10.1038/s43856-023-00421-7

18. Chen LC, Papandreou G, Schroff F, Adam H. Rethinking Atrous Convolution for Semantic Image Segmentation. Published online 2017. doi:10.48550/ARXIV.1706.05587

19. Liu Z, Mao H, Wu CY, Feichtenhofer C, Darrell T, Xie S. A ConvNet for the 2020s. Published online March 2, 2022. doi:10.48550/arXiv.2201.03545

20. Shen Y, Luo Y, Shen D, Ke J. RandStainNA: Learning Stain-Agnostic Features from Histology Slides by Bridging Stain Augmentation and Normalization. In: Vol 13432. ; 2022:212–221. doi:10.1007/978-3-031-16434-7_21

21. Hwang JE, Kim SS, Bang HJ, et al. Tumor Immune Microenvironment Biomarkers for Recurrence Prediction in Locally Advanced Rectal Cancer Patients after Neoadjuvant Chemoradiotherapy. Cancers. 2024;16(19):3353. doi:10.3390/cancers16193353

22. Fridman WH, Meylan M, Petitprez F, Sun CM, Italiano A, Sautès-Fridman C. B cells and tertiary lymphoid structures as determinants of tumour immune contexture and clinical outcome. Nat Rev Clin Oncol. 2022;19(7):441–457. doi:10.1038/s41571-022-00619-z

23. Trüb M, Zippelius A. Tertiary Lymphoid Structures as a Predictive Biomarker of Response to Cancer Immunotherapies. Front Immunol. 2021;12:674565. doi:10.3389/fimmu.2021.674565

24. Hao D, Han G, Sinjab A, et al. The Single-Cell Immunogenomic Landscape of B and Plasma Cells in Early-Stage Lung Adenocarcinoma. Cancer Discovery. 2022;12(11):2626–2645. doi:10.1158/2159-8290.CD-21-1658

25. Goff PH, Riolobos L, LaFleur BJ, et al. Neoadjuvant Therapy Induces a Potent Immune Response to Sarcoma, Dominated by Myeloid and B Cells. Clinical Cancer Research. 2022;28(8):1701–1711. doi:10.1158/1078-0432.CCR-21-4239

26. Thommen DS, Koelzer VH, Herzig P, et al. A transcriptionally and functionally distinct PD-1+ CD8+ T cell pool with predictive potential in non-small-cell lung cancer treated with PD-1 blockade. Nat Med. 2018;24(7):994–1004. doi:10.1038/s41591-018-0057-z

27. Patil NS, Nabet BY, Müller S, et al. Intratumoral plasma cells predict outcomes to PD-L1 blockade in non-small cell lung cancer. Cancer Cell. 2022;40(3):289–300.e4. doi:10.1016/j.ccell.2022.02.002

28. Barmpoutis P, Di Capite M, Kayhanian H, et al. Tertiary lymphoid structures (TLS) identification and density assessment on H&E-stained digital slides of lung cancer. Brcic L, ed. PLoS ONE. 2021;16(9):e0256907. doi:10.1371/journal.pone.0256907

29. Chen Z, Wang X, Jin Z, et al. Deep learning on tertiary lymphoid structures in hematoxylin-eosin predicts cancer prognosis and immunotherapy response. npj Precis Onc. 2024;8(1):73. doi:10.1038/s41698-024-00579-w

30. Rakaee M, Adib E, Ricciuti B, et al. Artificial intelligence in digital pathology approach identifies the predictive impact of tertiary lymphoid structures with immune-checkpoints therapy in NSCLC. JCO. 2022;40(16_suppl):9065–9065. doi:10.1200/JCO.2022.40.16_suppl.9065

31. Garassino MC, Gadgeel S, Novello S, et al. Associations of Tissue Tumor Mutational Burden and Mutational Status With Clinical Outcomes With Pembrolizumab Plus Chemotherapy Versus Chemotherapy For Metastatic NSCLC. JTO Clinical and Research Reports. 2023;4(1):100431. doi:10.1016/j.jtocrr.2022.100431

32. Mansfield AS, Aubry MC, Moser JC, et al. Temporal and spatial discordance of programmed cell death-ligand 1 expression and lymphocyte tumor infiltration between paired primary lesions and brain metastases in lung cancer. Annals of Oncology. 2016;27(10):1953–1958. doi:10.1093/annonc/mdw289

33. Zhang J, Dang F, Ren J, Wei W. Biochemical Aspects of PD-L1 Regulation in Cancer Immunotherapy. Trends in Biochemical Sciences. 2018;43(12):1014–1032. doi:10.1016/j.tibs.2018.09.004

34. Montfort A, Pearce O, Maniati E, et al. A Strong B-cell Response Is Part of the Immune Landscape in Human High-Grade Serous Ovarian Metastases. Clinical Cancer Research. 2017;23(1):250–262. doi:10.1158/1078-0432.CCR-16-0081

35. Germain C, Gnjatic S, Tamzalit F, et al. Presence of B Cells in Tertiary Lymphoid Structures Is Associated with a Protective Immunity in Patients with Lung Cancer. Am J Respir Crit Care Med. 2014;189(7):832–844. doi:10.1164/rccm.201309-1611OC

