## Supplementray_TLS for "Artificial Intelligence (AI)-Powered H&E Whole-Slide Image Analysis of Tertiary Lymphoid Structure (TLS) Independently Predicts Survival in Patients with Non-Small Cell Lung Cancer (NSCLC) Receiving Immunotherapy"

Supplementary Table 1. Mutational profiles of non-small cell lung cancer (NSCLC) samples in The Cancer Genome Atlas (TCGA) according to the presence of tertiary lymphoid structure (TLS) determined by AI analyzer

| **Characteristics** | **No. (%) (N = 910)** | | ***p*-value** |
|---|---|---|---|
| | **TLS (+) (N=776)** | **TLS (-) (N=134)** | |
| **Cancer type** | | | |
| LUAD | 383 (49.4%) | 74 (55.2%) | 0.246 |
| LUSC | 393 (50.6%) | 60 (44.8%) | |
| **EGFR** | | | |
| Mutated | 43 ( 5.5%) | 12 ( 9.0%) | 0.182 |
| WT | 733 (94.5%) | 122 (91.0%) | |
| **KRAS** | | | |
| Mutated | 121 (15.6%) | 27 (20.1%) | 0.233 |
| WT | 655 (84.4%) | 107 (79.9%) | |
| **BRAF** | | | |
| Mutated | 32 ( 4.1%) | 4 ( 3.0%) | 0.701 |
| WT | 744 (95.9%) | 130 (97.0%) | |
| **ERBB2** | | | |
| Mutated | 3 ( 0.4%) | 1 ( 0.7%) | 0.472 |
| WT | 773 (99.6%) | 133 (99.3%) | |
| **MET** | | | |
| Splicing variant | 4 ( 0.5%) | 2 ( 1.5%) | 0.217 |
| WT/other mutation | 772 (99.5%) | 132 (98.5%) | |
| **ALK** | | | |
| Translocation | 4 ( 0.5%) | 0 ( 0.0%) | 1 |
| WT | 772 (99.5%) | 134 (100.0%) | |
| **ROS1** | | | |
| Translocation | 4 ( 0.5%) | 2 ( 1.5%) | 0.217 |
| WT | 772 (99.5%) | 132 (98.5%) | |
| **RET** | | | |
| Translocation | 1 ( 0.1%) | 1 ( 0.7%) | 0.273 |
| WT | 775 (99.9%) | 133 (99.3%) | |
| **NTRK1** | | | |
| WT | 776 (100.0%) | 134 (100.0%) | NA |
| **NTRK2** | | | |
| Translocation | 0 ( 0.0%) | 1 ( 0.7%) | 0.147 |

| | | | |
|---|---|---|---|
| WT | 776 (100.0%) | 133 (99.3%) | |
| **NTRK3** | | | |
| WT | 776 (100.0%) | 134 (100.0%) | NA |

Abbreviations: LUAD, lung adenocarcinoma; LUSC, lung squamous cell carcinoma; WT, wild type; NA, not available.

Supplementary Table 2. Gene Set Enrichment Analysis (GSEA) results according to the MSigDB Hallmark gene sets and GO:BP.

| Pathway | *p*-value | $p_{adj}$ | ES | NES |
|---|---|---|---|---|
| **GO:BP pathway** | | | | |
| IMMUNOGLOBULIN PRODUCTION | 0.000 | 0.020 | 0.742 | 2.171 |
| KERATINIZATION | 0.000 | 0.020 | 0.768 | 2.120 |
| INTERMEDIATE FILAMENT ORGANIZATION | 0.000 | 0.020 | 0.724 | 1.977 |
| SPLICEOSOMAL TRI SNRNP COMPLEX ASSEMBLY | 0.000 | 0.021 | 0.822 | 1.970 |
| NUCLEOSOME ORGANIZATION | 0.000 | 0.020 | 0.651 | 1.863 |
| B CELL RECEPTOR SIGNALING PATHWAY | 0.000 | 0.020 | 0.676 | 1.847 |
| KILLING OF CELLS OF ANOTHER ORGANISM | 0.000 | 0.020 | 0.701 | 1.826 |
| XENOBIOTIC METABOLIC PROCESS | 0.000 | 0.020 | 0.641 | 1.823 |
| ANTIMICROBIAL HUMORAL RESPONSE | 0.000 | 0.032 | 0.603 | 1.717 |
| DIGESTION | 0.000 | 0.020 | 0.593 | 1.700 |
| TERPENOID METABOLIC PROCESS | 0.000 | 0.032 | 0.597 | 1.668 |
| NEGATIVE REGULATION OF ENDOPEPTIDASE ACTIVITY | 0.000 | 0.020 | 0.569 | 1.641 |
| **HALLMARK pathway** | | | | |
| XENOBIOTIC METABOLISM | 0.002 | 0.042 | 0.489 | 1.431 |
| KRAS SIGNALING DN | 0.003 | 0.042 | 0.488 | 1.428 |
| APICAL JUNCTION | 0.008 | 0.042 | -0.326 | -1.388 |
| KRAS SIGNALING UP | 0.008 | 0.042 | -0.332 | -1.419 |
| G2M CHECKPOINT | 0.008 | 0.042 | -0.397 | -1.690 |
| TNFA SIGNALING VIA NFKB | 0.008 | 0.042 | -0.397 | -1.692 |
| MITOTIC SPINDLE | 0.008 | 0.042 | -0.451 | -1.920 |
| UV RESPONSE DN | 0.004 | 0.042 | -0.484 | -1.999 |
| TGF BETA SIGNALING | 0.001 | 0.041 | -0.562 | -2.011 |
| EPITHELIAL MESENCHYMAL TRANSITION | 0.008 | 0.042 | -0.519 | -2.218 |

Abbreviations: ES, enrichment score; NES, normalized enrichment score; $p_{adj,}$ adjusted p value;

Supplementary Table 3. Patient characteristics with TLS assessed by a pathologist.

| **Characteristic** | **Overall**<br>N = 102[1] | **TLS negative**<br>N = 79[1] | **TLS positive**<br>N = 23[1] | ***p*-value**[2] |
|---|---|---|---|---|
| Age | 68 (59, 75) | 67 (59, 74) | 69 (60, 78) | 0.51 |
| Sex | | | | 0.25 |
| Female | 55 (54%) | 45 (57%) | 10 (43%) | |
| Male | 47 (46%) | 34 (43%) | 13 (57%) | |
| Smoking history | | | | 1.00 |
| Ever | 86 (84%) | 66 (84%) | 20 (87%) | |
| Never | 16 (16%) | 13 (16%) | 3 (13%) | |
| ECOG | | | | 0.23 |
| 0-1 | 82 (80%) | 61 (77%) | 21 (91%) | |
| 2-3 | 20 (20%) | 18 (23%) | 2 (8.7%) | |
| Stage | | | | 1.00 |
| III | 10 (9.8%) | 8 (10%) | 2 (8.7%) | |
| IV | 92 (90%) | 71 (90%) | 21 (91%) | |
| Pathology | | | | 0.64 |
| Non-squamous | 79 (77%) | 62 (78%) | 17 (74%) | |
| Squamous | 23 (23%) | 17 (22%) | 6 (26%) | |
| PDL-1 | | | | 0.17 |
| <1% | 28 (33%) | 22 (34%) | 6 (29%) | |
| ≥50% | 19 (22%) | 17 (26%) | 2 (9.5%) | |
| 1-49% | 39 (45%) | 26 (40%) | 13 (62%) | |
| Unknown | 16 | 14 | 2 | |
| TMB | | | | 1.00 |
| TMB-high (≥10/Mb) | 14 (24%) | 10 (23%) | 4 (27%) | |
| TMB-low (<10/Mb) | 44 (76%) | 33 (77%) | 11 (73%) | |
| Unknown | 44 | 36 | 8 | |
| Regimen | | | | <0.01 |
| ICI chemotherapy combination | 52 (51%) | 33 (42%) | 19 (83%) | |
| ICI monotherapy | 50 (49%) | 46 (58%) | 4 (17%) | |
| Treatment line | | | | 0.04 |
| ≥Second-line | 46 (45%) | 40 (51%) | 6 (26%) | |
| First-line | 56 (55%) | 39 (49%) | 17 (74%) | |
| Tissue harvest site | | | | 0.06 |
| Distant metastasis | 29 (28%) | 22 (28%) | 7 (30%) | |

| | | | | |
|---|---|---|---|---|
| Lymph node metastasis | 22 (22%) | 21 (27%) | 1 (4.3%) | |
| Primary tumor | 51 (50%) | 36 (46%) | 15 (65%) | |
| Specimen_type | | | | 0.12 |
| Biopsy | 63 (62%) | 52 (66%) | 11 (48%) | |
| Surgery | 39 (38%) | 27 (34%) | 12 (52%) | |
| Immune phenotype | | | | 0.19 |
| Inflamed | 16 (16%) | 10 (13%) | 6 (26%) | |
| Non-inflamed | 85 (84%) | 68 (87%) | 17 (74%) | |
| Unknown | 1 | 1 | 0 | |
| Inflamed score | | | | <0.01 |
| Above median | 50 (50%) | 33 (42%) | 17 (74%) | |
| Below median | 51 (50%) | 45 (58%) | 6 (26%) | |
| Unknown | 1 | 1 | 0 | |

[1] Median (Q1, Q3); n (%)

[2] Wilcoxon rank sum test; Pearson’s Chi-squared test; Fisher’s exact test

Supplementary Table 4. Multiple univariate survival analyses of PFS (A) and OS (B) following ICI treatment stratified by baseline patient characteristics and the presence of TLS as assessed by a pathologist and an AI analyzer. AI, artificial intelligence; CI, confidence interval; ECOG, Eastern Cooperative Oncology Group Performance Status; HR, hazard ratio; ICI, immune checkpoint inhibitor; OS, overall survival; PD-L1, programmed death ligand-1; PFS, progression-free survival; TLS, tertiary lymphoid structure.

(A) PFS

| **Variable** | | **HR (95% CI)** | ***p*-value** |
|---|---|---|---|
| TLS assessed by AI | TLS Positive (vs Negative) | 0.49 (0.29–0.83) | <0.01 |
| TLS assessed by a pathologist | TLS Positive (vs Negative) | 0.45 (0.25–0.82) | <0.01 |
| Age | | 1.00 (0.98–1.03) | 0.71 |
| Sex | Female (vs Male) | 0.74 (0.48–1.15) | 0.19 |
| ECOG | 0-1 (vs 2-3) | 0.86 (0.49–1.48) | 0.58 |
| Smoking history | Never (vs Ever) | 1.48 (0.83–2.64) | 0.18 |
| Pathology | Squamous (vs Non-squamous) | 0.92 (0.55–1.54) | 0.75 |
| Stage | III (vs IV) | 0.80 (0.37–1.74) | 0.57 |
| Regimen | ICI chemotherapy combination (vs ICI monotherapy) | 0.83 (0.53–1.28) | 0.40 |
| Line of treatment | First line (vs ≥ Second-line) | 0.59 (0.38–0.92) | 0.02 |
| PD-L1 expression level | 1-49% (< 1%) | 1.29 (0.74–2.26) | 0.37 |
| | ≥50% (< 1%) | 1.04 (0.53–2.01) | 0.91 |
| Immune phenotype | Inflamed (vs Non-inflamed) | 0.56 (0.29–1.06) | 0.07 |
| Inflamed score | Above median (vs Below median) | 0.65 (0.42–1.02) | 0.06 |
| Inflamed score (numeric) | | 0.99 (0.97–1.00) | 0.04 |

(B) OS

| **Variable** | | **HR (95% CI)** | ***p*-value** |
|---|---|---|---|
| TLS assessed by AI | TLS Positive (vs Negative) | 0.55 (0.33–0.92) | 0.02 |
| TLS assessed by a pathologist | TLS Positive (vs Negative) | 0.43 (0.23–0.78) | <0.01 |
| Age | | 1.00 (0.98–1.02) | 0.92 |
| Sex | Female (vs Male) | 0.66 (0.42–1.03) | 0.07 |
| ECOG | 0-1 (vs 2-3) | 0.53 (0.31–0.91) | 0.02 |
| Smoking history | Never (vs Ever) | 1.02 (0.56–1.86) | 0.95 |
| Pathology | Squamous (vs Non-squamous) | 0.88 (0.51–1.51) | 0.64 |
| Stage | III (vs IV) | 0.82 (0.35–1.88) | 0.64 |
| Regimen | ICI chemotherapy combination (vs ICI monotherapy) | 0.69 (0.44–1.08) | 0.11 |
| Line of treatment | First line (vs ≥ Second-line) | 0.52 (0.33–0.83) | <0.01 |
| PD-L1 expression level | 1-49% (< 1%) | 1.18 (0.67–2.10) | 0.57 |
| | ≥50% (< 1%) | 1.29 (0.66–2.51) | 0.45 |
| Immune phenotype | Inflamed (vs Non-inflamed) | 0.63 (0.34–1.18) | 0.15 |
| Inflamed score | Above median (vs Below median) | 0.72 (0.46–1.13) | 0.15 |
| Inflamed score (numeric) | | 0.99 (0.98–1.00) | 0.10 |

Supplementary Table 5. Multivariable survival analyses of PFS (A) and OS (B) following ICI treatment stratified by baseline patient characteristics excluding TMB and the presence of TLS as assessed by AI analyzer. Multivariable survival analyses of PFS (C) and OS (D) following ICI treatment stratified by baseline patient characteristics excluding TMB and the presence of TLS as assessed by a pathologist. AI, artificial intelligence; CI, confidence interval;ECOG, Eastern Cooperative Oncology Group Performance Status; HR, hazard ratio; ICI, immune checkpoint inhibitor; OS, overall survival; PD-L1, programmed death ligand-1; PFS, progression-free survival; TLS, tertiary lymphoid structure.

(A) PFS

| **Variable** | | **HR (95% CI)** | ***p*-value** |
|---|---|---|---|
| TLS assessed by AI | TLS Positive (vs Negative) | 0.40 (0.21–0.77) | <0.01 |
| Age | | 1.02 (0.99–1.05) | 0.19 |
| Sex | Female (vs Male) | 0.47 (0.26–0.84) | 0.01 |
| ECOG | 0-1 (vs 2-3) | 0.68 (0.36–1.29) | 0.24 |
| Smoking history | Never (vs Ever) | 1.52 (0.68–3.38) | 0.31 |
| Pathology | Squamous (vs Non-squamous) | 1.05 (0.54–2.03) | 0.90 |
| Stage | III (vs IV) | 0.68 (0.26–1.80) | 0.44 |
| Regimen | ICI chemotherapy combination (vs ICI monotherapy) | 1.19 (0.68–2.10) | 0.55 |
| Line of treatment | First line (vs ≥ Second-line) | 0.50 (0.28–0.90) | 0.02 |
| PD-L1 expression level | 1-49% (< 1%) | 1.29 (0.69–2.41) | 0.42 |
| | ≥50% (< 1%) | 1.25 (0.60–2.59) | 0.55 |
| Inflamed score (numeric) | | 1.00 (0.98–1.01) | 0.63 |

(B) OS

| **Variable** | | **HR (95% CI)** | ***p*-value** |
|---|---|---|---|
| TLS assessed by AI | TLS Positive (vs Negative) | 0.52 (0.28–0.99) | 0.05 |
| Age | | 1.01 (0.98–1.04) | 0.47 |
| Sex | Female (vs Male) | 0.41 (0.23–0.74) | <0.01 |
| ECOG | 0-1 (vs 2-3) | 0.41 (0.21–0.78) | <0.01 |
| Smoking history | Never (vs Ever) | 1.13 (0.48–2.65) | 0.78 |
| Pathology | Squamous (vs Non-squamous) | 1.05 (0.53–2.10) | 0.88 |
| Stage | III (vs IV) | 0.82 (0.28–2.35) | 0.71 |
| Regimen | ICI chemotherapy combination (vs ICI monotherapy) | 0.99 (0.54–1.81) | 0.97 |
| Line of treatment | First line (vs ≥ Second-line) | 0.41 (0.22–0.78) | <0.01 |
| PD-L1 expression level | 1-49% (< 1%) | 1.12 (0.60–2.10) | 0.72 |
| | ≥50% (< 1%) | 1.63 (0.75–3.53) | 0.22 |
| Inflamed score (numeric) | | 0.99 (0.98–1.01) | 0.33 |

(C) PFS

| Variable | | HR (95% CI) | *p*-value |
|---|---|---|---|
| TLS assessed by a pathologist | TLS Positive (vs Negative) | 0.36 (0.18–0.73) | <0.01 |
| Age | | 1.02 (1.00–1.05) | 0.10 |
| Sex | Female (vs Male) | 0.44 (0.24–0.80) | <0.01 |
| ECOG | 0-1 (vs 2-3) | 0.78 (0.41–1.50) | 0.46 |
| Smoking history | Never (vs Ever) | 1.75 (0.78–3.92) | 0.17 |
| Pathology | Squamous (vs Non-squamous) | 0.92 (0.47–1.81) | 0.81 |
| Stage | III (vs IV) | 0.82 (0.31–2.13) | 0.68 |
| Regimen | ICI chemotherapy combination (vs ICI monotherapy) | 1.18 (0.67–2.08) | 0.57 |
| Line of treatment | First line (vs ≥ Second-line) | 0.48 (0.27–0.88) | 0.02 |
| PD-L1 expression level | 1-49% (< 1%) | 1.25 (0.67–2.33) | 0.48 |
| | ≥50% (< 1%) | 1.03 (0.49–2.16) | 0.94 |
| Inflamed score (numeric) | | 0.99 (0.98–1.01) | 0.44 |

(D) OS

| Variable | | HR (95% CI) | *p*-value |
|---|---|---|---|
| TLS assessed by a pathologist | TLS Positive (vs Negative) | 0.42 (0.21–0.85) | 0.02 |
| Age | | 1.01 (0.99–1.04) | 0.32 |
| Sex | Female (vs Male) | 0.37 (0.21–0.68) | <0.01 |
| ECOG | 0-1 (vs 2-3) | 0.44 (0.22–0.86) | 0.02 |
| Smoking history | Never (vs Ever) | 1.26 (0.54–2.97) | 0.59 |
| Pathology | Squamous (vs Non-squamous) | 0.97 (0.49–1.94) | 0.94 |
| Stage | III (vs IV) | 0.90 (0.32–2.55) | 0.85 |
| Regimen | ICI chemotherapy combination (vs ICI monotherapy) | 0.98 (0.54–1.79) | 0.94 |
| Line of treatment | First line (vs ≥ Second-line) | 0.39 (0.21–0.74) | <0.01 |
| PD-L1 expression level | 1-49% (< 1%) | 1.12 (0.60–2.09) | 0.73 |
| | ≥50% (< 1%) | 1.44 (0.67–3.11) | 0.35 |
| Inflamed score (numeric) | | 0.99 (0.98–1.01) | 0.30 |

Supplementary Figure 1. Flow chart of retrospective patient selection for the study.

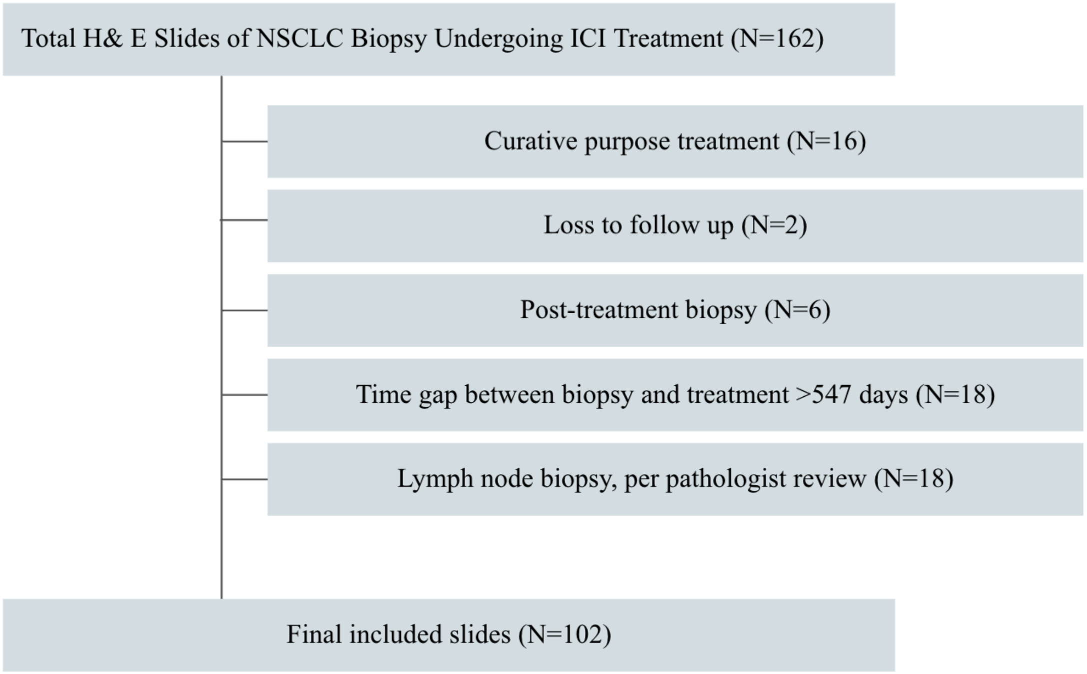

Supplementary Figure 2. Representative example of a false-positive tertiary lymphoid structures (TLS) detection by the artificial intelligence (AI) analyzer. The figure presents a whole slide image (WSI) of a representative case where the AI analyzer incorrectly identified TLS. On the left, the WSI is displayed at low magnification, with areas falsely detected as TLS highlighted in orange. The right side of the figure shows a high-magnification view.

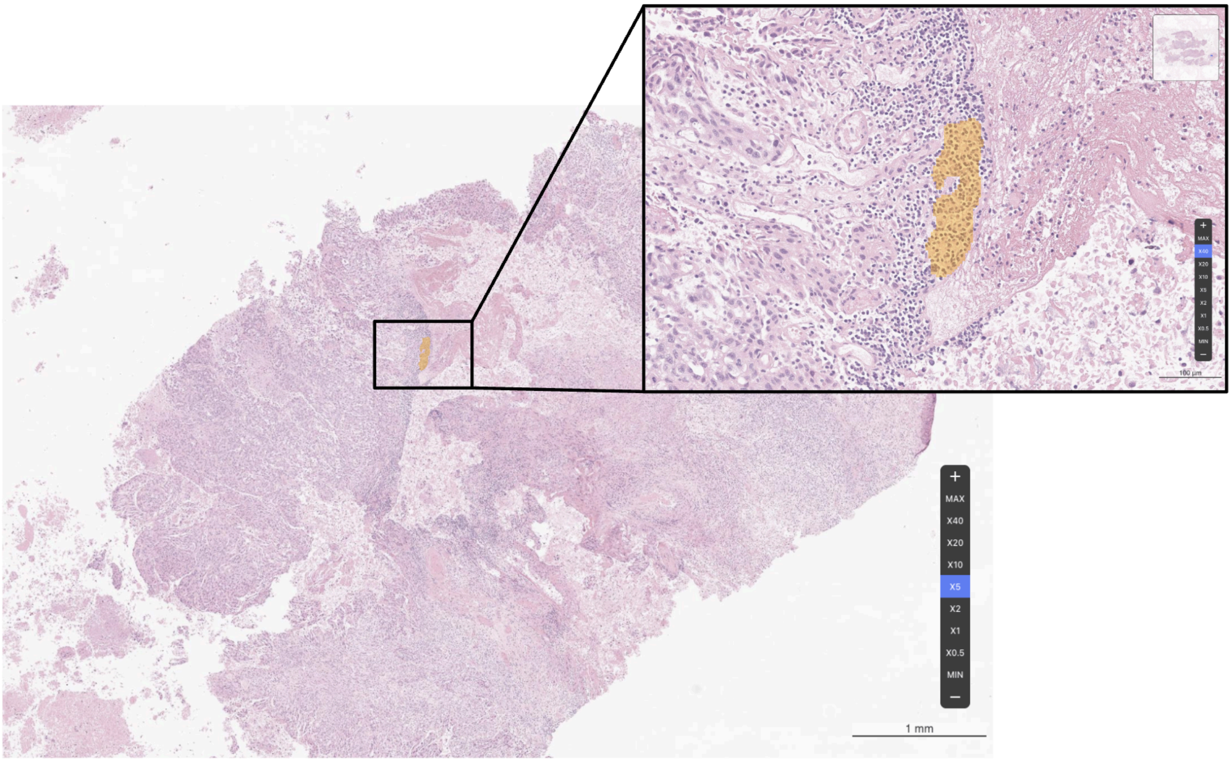

Supplementary Figure 3. (A) Kaplan-Meier survival analysis of PFS (left) and OS (right) following ICI treatment based on TLS presence categorized by AI and pathologist agreement. P-values were calculated using a two-sided log-rank test. The Cox proportional hazards model was used for calculation of HRs and corresponding 95% CIs. (B) Table summarizing the result of survival analysis following ICI treatment based on the presence of a pathologist vs AI analyzer. AI, artificial intelligence; CI, confidence interval; HR, hazard ratio; ICI, immune checkpoint inhibitor; NR, not reached; ORR, objective response rate; OS, overall survival; PD-L1, programmed death ligand-1; PFS, progression-free survival; TLS, tertiary lymphoid structure.

(A)

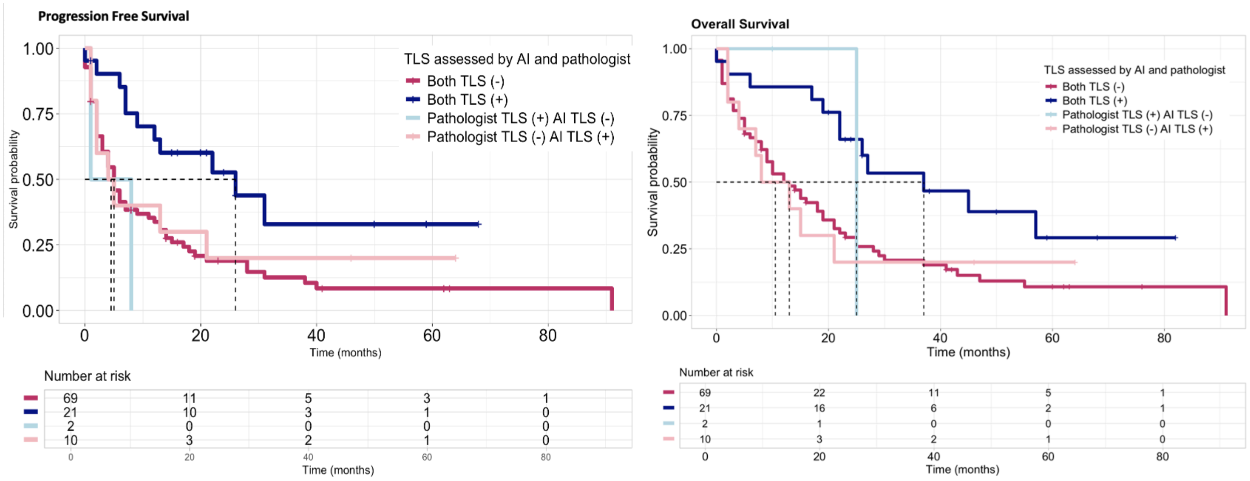


(B)

| | | PFS | | OS | |
|---|---|---|---|---|---|
| | | **HR (95% CI)** | **p-value** | **HR (95% CI)** | **p-value** |
| **Pathologist** | TLS (-) (n = 79) | Reference | **<0.01*** | Reference | **<0.01*** |
| | TLS (+) (n = 23) | 0.45 (0.25-0.82) | | 0.43 (0.23-0.78) | |
| **AI analyzer** | TLS (-) (n = 71) | Reference | **<0.01*** | Reference | **0.02*** |
| | TLS (+) (n = 31) | 0.49 (0.29-0.82) | | 0.55 (0.33-0.92) | |
| **Pathologist + AI analyzer** | Both TLS (-) (n=69) | Reference | | Reference | |
| | Pathologist TLS (+), AI TLS (-) (n=2) | 1.82 (0.44-7.50) | 0.41 | 0.53 (0.07-3.84) | 0.53 |
| | Pathologist TLS (-), AI TLS (+) (n=10) | 0.83 (0.39-1.73) | 0.61 | 0.98 (0.47-2.06) | 0.96 |
| | Both TLS (+) (n=21) | 0.39 (0.20-0.74) | **<0.01*** | 0.42 (0.22-0.78) | **<0.01*** |

Supplementary Figure 4. (A) Kaplan-Meier survival analysis of PFS (left) and OS (right) following ICI monotherapy based on the presence of TLS assessed by a pathologist. (B) Kaplan-Meier survival analysis of PFS (left) and OS (right) following ICI and chemotherapy combination based on the presence of TLS assessed by a pathologist. P-values were calculated using a two-sided log-rank test. The Cox proportional hazards model was used for calculation of HRs and corresponding 95% CIs. AI, artificial intelligence; CI, confidence interval; HR, hazard ratio; ICI, immune checkpoint inhibitor; NR, not reached; ORR, objective response rate; OS, overall survival; PFS, progression-free survival; TLS, tertiary lymphoid structure.

(A) ICI monotherapy

| Group | N | ORR (%) | Median PFS (months, 95% CI) | HR (95% CI) | P-value |
|---|---|---|---|---|---|
| TLS negative | 46 | 10.9 | 4 (2 - 13) | Ref. | Ref. |
| TLS positive | 4 | 25.0 | NR (NR - NR) | 0 (0 - Inf) | <0.01 |

| Group | Median OS (months, 95% CI) | HR (95% CI) | P-value |
|---|---|---|---|
| TLS negative | 12 (6 - 21) | Ref. | Ref. |
| TLS positive | NR (NR – NR) | 0 (0 - Inf) | <0.01 |

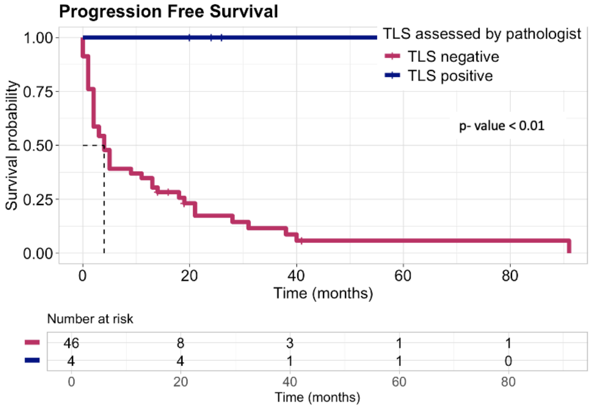


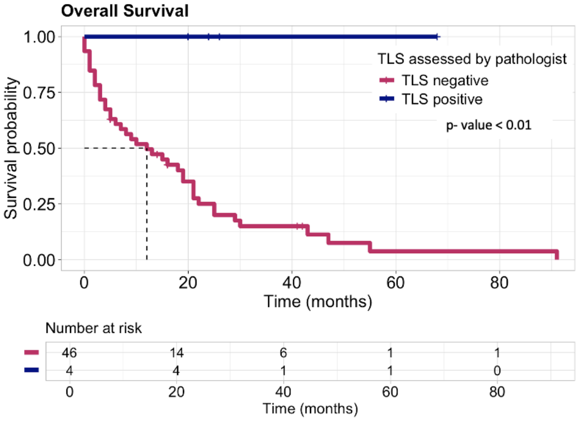


(B) ICI and chemotherapy combination

| Group | N | ORR (%) | Median PFS (months, 95% CI) | HR (95% CI) | P-value |
|---|---|---|---|---|---|
| TLS negative | 33 | 27.3 | 6 (4 - 14) | Ref. | Ref. |
| TLS positive | 19 | 15.8 | 13 (7 - NR) | 0.65 (0.33 - 1.26) | 0.21 |

| Group | Median OS (months, 95% CI) | HR (95% CI) | P-value |
|---|---|---|---|
| TLS negative | 13 (9- 28) | Ref. | Ref. |
| TLS positive | 26 (22 – NR) | 0.64 (0.33 - 1.26) | 0.19 |

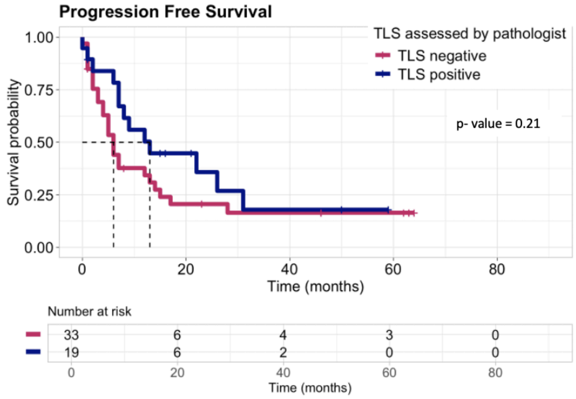


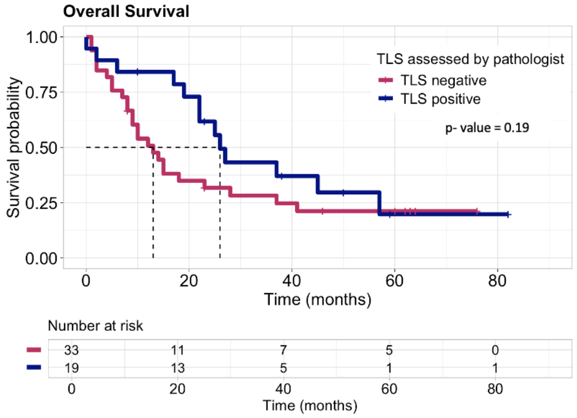

Supplementary Figure 5. Kaplan-Meier survival analyses of PFS (left) and OS (right) following ICI treatment as a first-line treatment, stratified by the presence of TLS as assessed by (A) a pathologist and (B) an AI analyzer. Kaplan-Meier survival analyses of PFS (left) and OS (right) following ICI treatment as a second-line or beyond treatment, stratified by the presence of TLS as assessed by (C) a pathologist and (D) an AI analyzer. P-values were calculated using a two-sided log-rank test. The Cox proportional hazards model was used for calculation of HRs and corresponding 95% CIs. AI, artificial intelligence; CI, confidence interval; HR, hazard ratio; ICI, immune checkpoint inhibitor; NR, not reached; ORR, objective response rate; OS, overall survival; PD-L1, programmed death ligand-1; PFS, progression-free survival; TLS, tertiary lymphoid structure.

(A) First-line

| Group | N | Median PFS (months, 95% CI) | HR (95% CI) | P-value |
|---|---|---|---|---|
| TLS negative | 39 | 11 (5 - 21) | Ref. | Ref. |
| TLS positive | 17 | 22 (8 – NR) | 0.60 (0.29 – 1.22) | 0.16 |

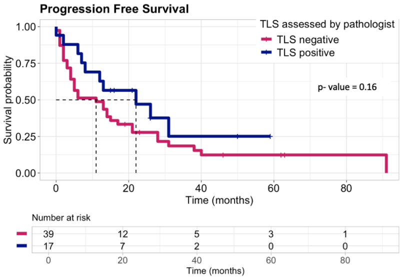


| Group | Median OS (months, 95% CI) | HR (95% CI) | P-value |
|---|---|---|---|
| TLS negative | 18 (13 - 41) | Ref. | Ref. |
| TLS positive | 37 (22 – NR) | 0.61 (0.30 – 1.24) | 0.17 |

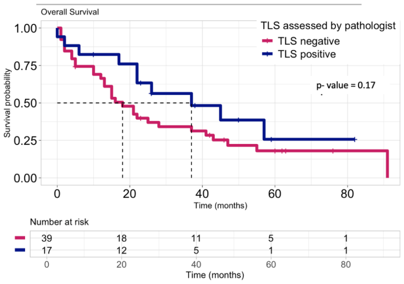


(B) First-line

| Group | N | Median PFS (months, 95% CI) | HR (95% CI) | P-value |
|---|---|---|---|---|
| TLS negative | 35 | 8 (5 – 21) | Ref. | Ref. |
| TLS positive | 21 | 21 (12 – NR) | 0.62 (0.33 – 1.20) | 0.15 |

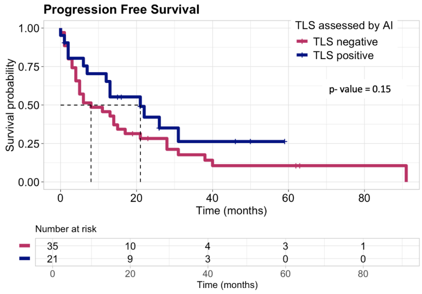


| Group | Median OS (months, 95% CI) | HR (95% CI) | P-value |
|---|---|---|---|
| TLS negative | 18 (13 - 43) | Ref. | Ref. |
| TLS positive | 26 (17 - NR) | 0.76 (0.39 – 1.45) | 0.40 |

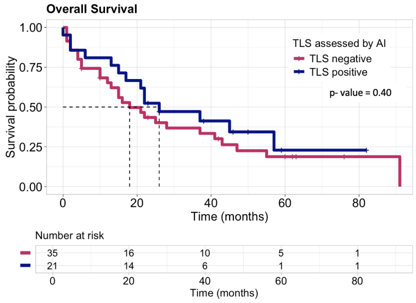


(C) ≥ Second-line

| Group | N | Median PFS (months, 95% CI) | HR (95% CI) | P-value |
|---|---|---|---|---|
| TLS negative | 40 | 3 (2 – 6) | Ref. | Ref. |
| TLS positive | 6 | 9 (7 – NR) | 0.31 (0.09 – 1.03) | 0.04 |

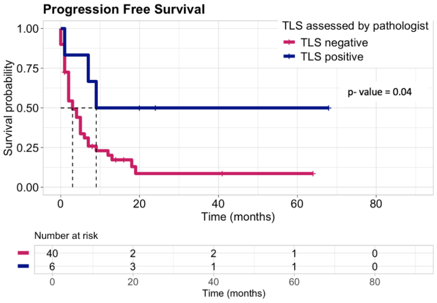


| Group | Median OS (months, 95% CI) | HR (95% CI) | P-value |
|---|---|---|---|
| TLS negative | 8 (5 – 18) | Ref. | Ref. |
| TLS positive | 27 (25 – NA) | 0.26 (0.08 – 0.85) | 0.02 |

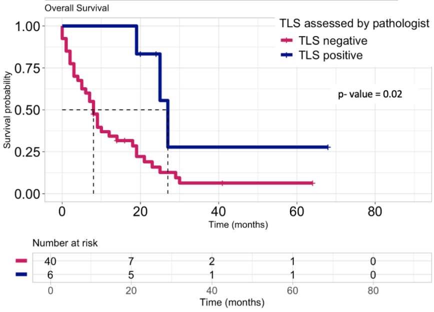


(D) ≥ Second-line

| Group | N | Median PFS (months, 95% CI) | HR (95% CI) | P-value |
|---|---|---|---|---|
| TLS negative | 36 | 3 (2 – 7) | Ref. | Ref. |
| TLS positive | 10 | 8 (4 – NR) | 0.41 (0.17 – 0.99) | 0.05 |

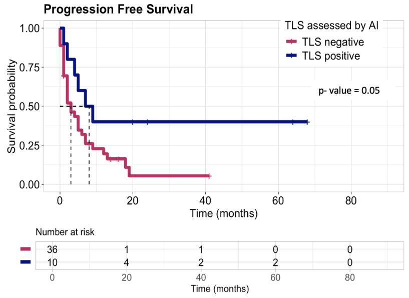


| Group | Median OS (months, 95% CI) | HR (95% CI) | P-value |
|---|---|---|---|
| TLS negative | 9 (5 – 19) | Ref. | Ref. |
| TLS positive | 23 (7 – NR) | 0.43 (0.18 – 1.03) | 0.05 |

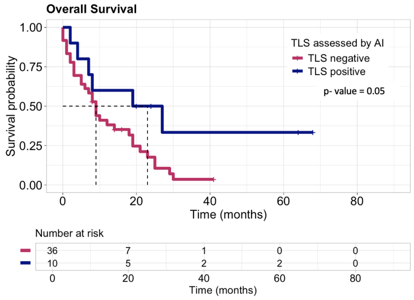

Supplementary Figure 6. Kaplan-Meier survival analysis of PFS (left) and OS (right) following ICI treatment in PD-L1 positive ( ≥1%) patients based on the presence of TLS assessed by (A) a pathologist, and (B) AI analyzer. Kaplan-Meier survival analysis of PFS (left) and OS (right) following ICI treatment in PD-L1 negative (<1%) patients based on the presence of TLS assessed by (C) a pathologist, and (D) AI analyzer. P-values were calculated using a two-sided log-rank test. The Cox proportional hazards model was used for calculation of HRs and corresponding 95% CIs. AI, artificial intelligence; CI, confidence interval; HR, hazard ratio; ICI, immune checkpoint inhibitor; NR, not reached; ORR, objective response rate; OS, overall survival; PD-L1, programmed death ligand-1; PFS, progression-free survival; TLS, tertiary lymphoid structure.

(A) PD-L1 positive ( ≥1%)

| Group | N | ORR (%) | Median PFS (months, 95% CI) | HR (95% CI) | P-value |
|---|---|---|---|---|---|
| TLS negative | 43 | 20.9 | 5 (4 – 15) | Ref. | Ref. |
| TLS positive | 15 | 20.0 | 13 (7 – NR) | 0.48 (0.23 – 0.98) | 0.05 |

| Group | Median OS (months, 95% CI) | HR (95% CI) | P-value |
|---|---|---|---|
| TLS negative | 13 (8 - 21) | Ref. | Ref. |
| TLS positive | 37 (27 – NR) | 0.34 (0.15 - 0.77) | < 0.01 |

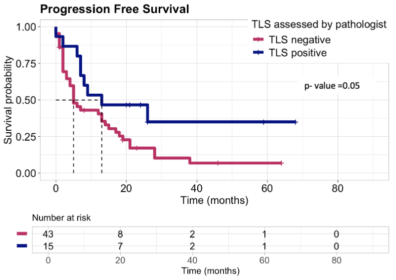


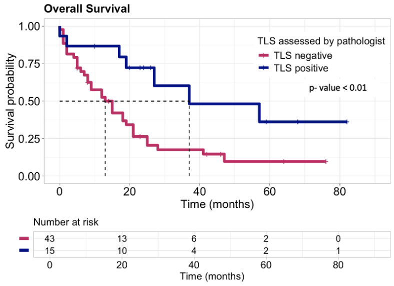


(B) PD-L1 positive ( ≥1%)

| Group | N | ORR (%) | Median PFS (months, 95% CI) | HR (95% CI) | P-value |
|---|---|---|---|---|---|
| TLS negative | 37 | 18.9 | 5 (4 – 14) | Ref. | Ref. |
| TLS positive | 21 | 23.8 | 13 (7 – NR) | 0.43 (0.22 – 0.83) | <0.01 |

| Group | Median OS (months, 95% CI) | HR (95% CI) | P-value |
|---|---|---|---|
| TLS negative | 12 (8 – 21) | Ref. | Ref. |
| TLS positive | 27 (17 – NR) | 0.40 (0.20 – 0.80) | <0.01 |

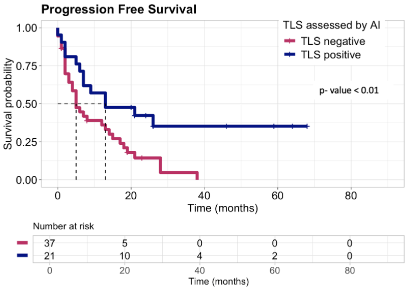


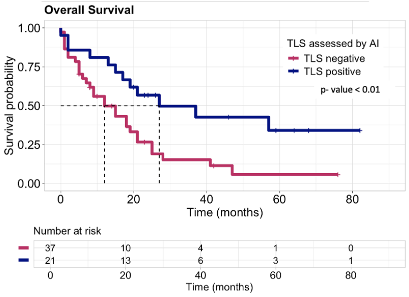


(C) PD-L1 negative (<1%)

| Group | N | ORR (%) | Median PFS (months, 95% CI) | HR (95% CI) | P-value |
|---|---|---|---|---|---|
| TLS negative | 22 | 9.1 | 6 (3 – 31) | Ref. | Ref. |
| TLS positive | 6 | 16.7 | 31 (22 – NR) | 0.37 (0.11 – 1.26) | 0.09 |

| Group | Median OS (months, 95% CI) | HR (95% CI) | P-value |
|---|---|---|---|
| TLS negative | 18.5 (8 – 55) | Ref. | Ref. |
| TLS positive | 35.5 (22 – NR) | 0.63 (0.21 – 1.89) | 0.40 |

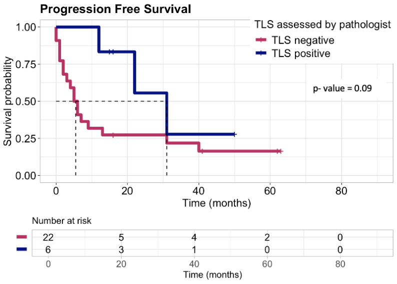


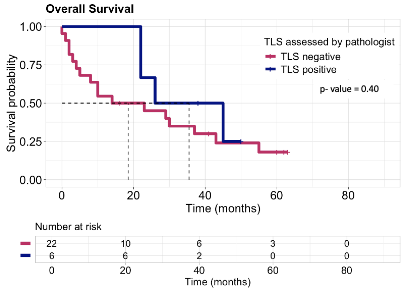


(D) PD-L1 negative (<1%)

| Group | N | ORR (%) | Median PFS (months, 95% CI) | HR (95% CI) | P-value |
|---|---|---|---|---|---|
| TLS negative | 21 | 9.5 | 6 (3 – 40) | Ref. | Ref. |
| TLS positive | 7 | 14.3 | 22 (12 – NR) | 0.50 (0.17 – 1.51) | 0.21 |

| Group | Median OS (months, 95% CI) | HR (95% CI) | P-value |
|---|---|---|---|
| TLS negative | 23 (8 – 55) | Ref. | Ref. |
| TLS positive | 26 (22 – NR) | 0.82 (0.30 – 2.25) | 0.68 |

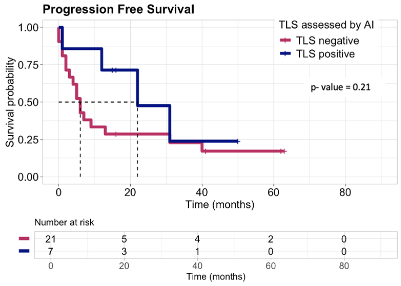


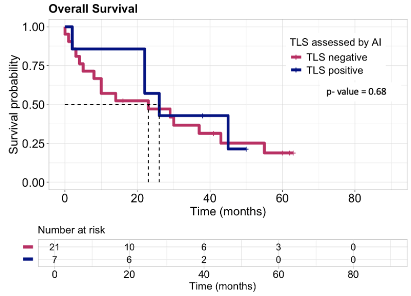

Supplementary Figure 7. Kaplan-Meier survival analysis of PFS (left) and OS (right) after ICI treatment according to (A) the proportion of TLS area to total tissue area calculated by AI analyzer and (B) the number of TLS clusters counted by AI analyzer. P-values were calculated using a two-sided log-rank test. The Cox proportional hazards model was used for calculation of HRs and corresponding 95% CIs. AI, artificial intelligence; CI, confidence interval; HR, hazard ratio; ICI, immune checkpoint inhibitor; NR, not reached; ORR, objective response rate; OS, overall survival; PFS, progression-free survival; TLS, tertiary lymphoid structure; TLSP, tertiary lymphoid structure proportion.

(A)

| Group | N | Median PFS (months, 95% CI) | HR (95% CI) | P-value |
|---|---|---|---|---|
| TLSP Lower than median | 15 | NR (31 – NR) | Ref. | Ref. |
| TLSP Higher than median | 16 | NR (7 – NR) | 1.42 (0.56 – 3.60) | 0.96 |

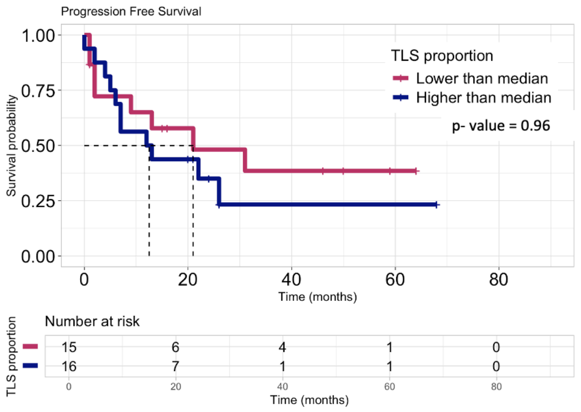


| Group | N | Median OS (months, 95% CI) | HR (95% CI) | P-value |
|---|---|---|---|---|
| TLSP Lower than median | 15 | 22 (12 – NR) | Ref. | Ref. |
| TLSP Higher than median | 16 | 26 (17 – NR) | 0.97 (0.40 – 2.35) | 0.96 |

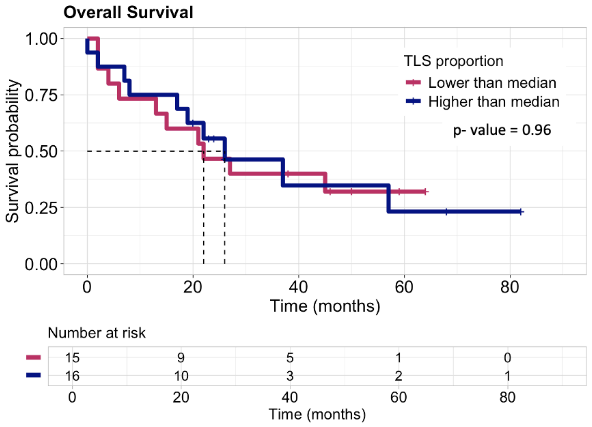


(B)

| Group | N | Median PFS (months, 95% CI) | HR (95% CI) | P-value |
|---|---|---|---|---|
| Number of clusters lower than median | 16 | 21 (6 – NR) | Ref. | Ref. |
| Number of clusters higher than median | 15 | 13 (7 – NR) | 0.96 (0.39 – 2.37) | 0.92 |

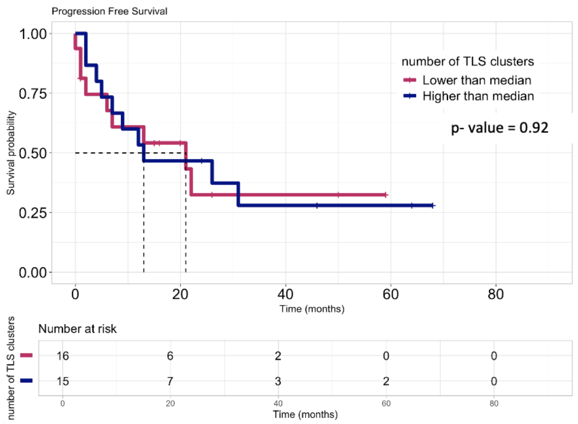


| Group | N | Median OS (months, 95% CI) | HR (95% CI) | P-value |
|---|---|---|---|---|
| Number of clusters lower than median | 16 | 21 (13 – NR) | Ref. | Ref. |
| Number of clusters higher than median | 15 | 36 (17 – NR) | 0.63 (0.26 – 1.55) | 0.32 |

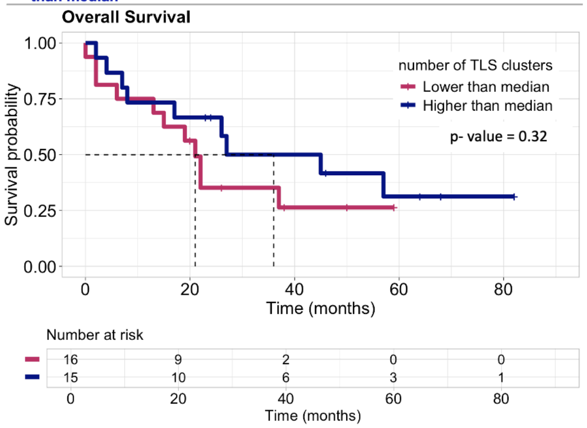

Supplementary Figure 8. Kaplan-Meier survival analysis of PFS (left) and OS (right) after ICI treatment according to (A) inflamed score and (B) immune phenotype. P-values were calculated using a two-sided log-rank test. The Cox proportional hazards model was used for calculation of HRs and corresponding 95% CIs. AI, artificial intelligence; CI, confidence interval; HR, hazard ratio; ICI, immune checkpoint inhibitor; NR, not reached; ORR, objective response rate; OS, overall survival; PFS, progression-free survival; TLS, tertiary lymphoid structure.

(A)

| Group | N | Median PFS (months, 95% CI) | HR (95% CI) | P-value |
|---|---|---|---|---|
| Inflamed score lower than median | 50 | 5 (3 – 13) | Ref. | Ref. |
| Inflamed score higher than median | 51 | 12 (6 – 28) | 0.65 (0.42 – 1.02) | 0.06 |

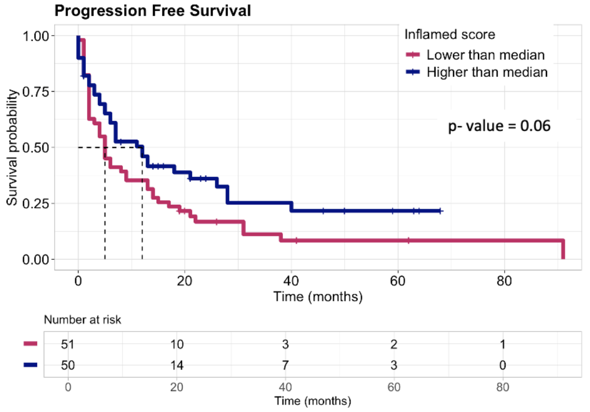


| Group | Median OS (months, 95% CI) | HR (95% CI) | P-value |
|---|---|---|---|
| Inflamed score lower than median | 13 (8 - 22) | Ref. | Ref. |
| Inflamed score higher than median | 19 (15 – 29) | 0.72 (0.46 – 1.13) | 0.15 |

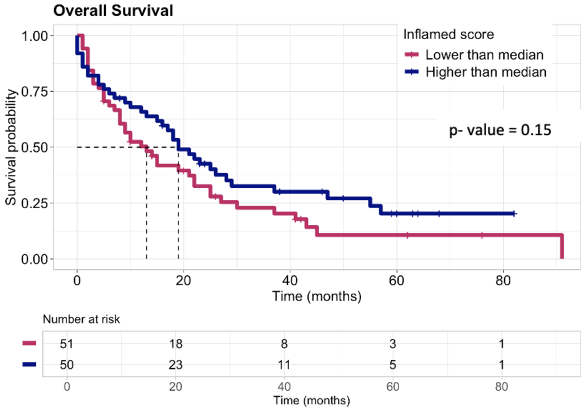


(B)

| Group | N | Median PFS (months, 95% CI) | HR (95% CI) | P-value |
|---|---|---|---|---|
| Non-inflamed | 85 | 6 (4 – 12) | Ref. | Ref. |
| Inflamed | 16 | 26 (5 – NR) | 0.56 (0.29 – 1.06) | 0.07 |

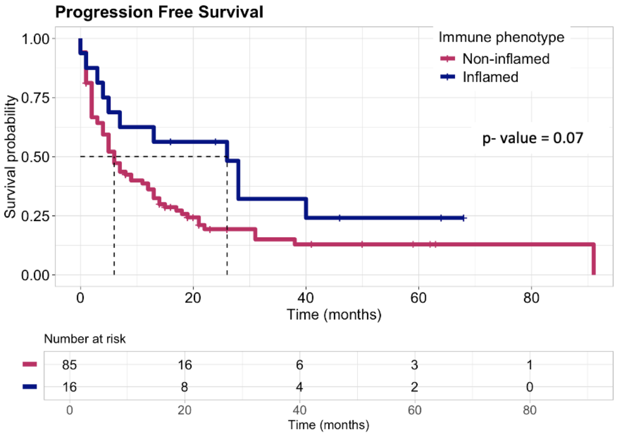


| Group | Median OS (months, 95% CI) | HR (95% CI) | P-value |
|---|---|---|---|
| Non-inflamed | 14 (10 - 22) | Ref. | Ref. |
| Inflamed | 28 (18 – NR) | 0.63 (0.34 – 1.18) | 0.15 |

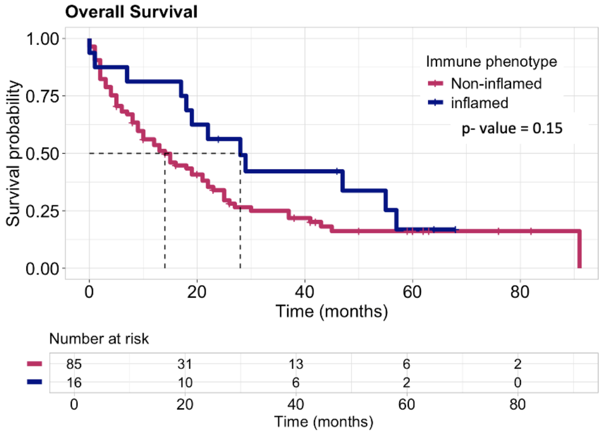

**Supplementary Method 1. Assessment of the AI-determined TLS and its association with genetic mutations and immunological characteristics in publicly available dataset**

The analyzer was applied to H&E-stained images of 924 NSCLC samples sourced from TCGA, comprising lung adenocarcinoma (LUAD, *N*=462) and lung squamous cell carcinoma (LUSC, *N*=462).

To investigate the relationship between TLS presence and genetic mutations, mutational data derived from whole exome sequencing for each case was obtained from cBioPortal (https://www.cbioportal.org/). This analysis focused on 11 driver mutations, including *EGFR*, *KRAS*, *BRAF*, *ERBB2* mutations; *MET* splicing variants; and translocations in *ROS1*, *ALK*, *RET*, and *NTRK 1/2/3*. The frequencies of these mutations were compared between TLS(+) and TLS(-) groups.

To assess the link between TLS presence and immunological characteristics, we retrieved gene expression profiles and clinical data for each case from TCGA. Immune cell infiltration levels were determined using CIBERSORT, which predicts the proportion of 22 tumor-infiltrating immune cells based on the relative expression of 547 genes. These cell types included seven T cell subsets, naive and memory B cells, plasma cells, NK cells, and myeloid subsets. Fold-change (fc) values and corresponding *p*-values were calculated for each immune cell type to compare TLS(+) and TLS(-) groups.

Differential gene expression analysis was performed on TCGA's RNA sequencing data using the DESeq2 R package (version 1.38.3). Genes exhibiting a |log2-fold change| > 1 and an adjusted *p*-value ($p_{adj}$) < 0.05 (Benjamini-Hochberg correction) were considered differentially expressed between the TLS(+) and TLS(-) groups.

Lastly, gene set enrichment analysis (GSEA) was conducted using the Fgsea R package (version 1.24.0) to identify differences in pathway activity between TLS(+) and TLS(-) groups. Using differentially expressed genes, we analyzed enrichment in Hallmark pathways or biological process components of gene ontology (C5 GO:BP) from the Human Molecular Signatures Database (MSigDB, https://www.gsea-msigdb.org/). Pathways with $P_{adj} < 0.05$ were considered significantly associated with TLS presence.
